# What has brain diffusion MRI taught us about chronic pain: a narrative review

**DOI:** 10.1101/2023.03.03.23286579

**Authors:** Paul Bautin, Marc-Antoine Fortier, Monica Sean, Graham Little, Marylie Martel, Maxime Descoteaux, Guillaume Léonard, Pascal Tétreault

## Abstract

Chronic pain is a pervasive and debilitating condition with increasing implications for public health, affecting millions of individuals worldwide. Despite its high prevalence, the underlying neural mechanisms and pathophysiology remain only partly understood. Since its introduction 35 years ago, brain diffusion MRI has emerged as a powerful tool to investigate changes in white matter microstructure and connectivity associated with chronic pain. This review synthesizes findings from 58 articles that constitute the current research landscape, covering methodologies and key discoveries.

We discuss the evidence supporting the role of altered white matter microstructure and connectivity in chronic pain conditions, highlighting the importance of studying multiple chronic pain syndromes to identify common neurobiological pathways. We also explore the prospective clinical utility of diffusion MRI, such as its role in identifying diagnostic, prognostic, and therapeutic biomarkers.

Further, we address shortcomings and challenges associated with brain diffusion MRI in chronic pain studies, emphasizing the need for the harmonization of data acquisition and analysis methods. We conclude by highlighting emerging approaches and prospective avenues in the field that may provide new insights into the pathophysiology of chronic pain and potential new therapeutic targets.

Due to the limited current body of research and unidentified targeted therapeutic strategies, we are forced to conclude that further research is required. However, we believe that brain diffusion MRI presents a promising opportunity for enhancing our understanding of chronic pain and improving clinical outcomes.

## Introduction

Over the past 25 years, significant progress has been made in the understanding of chronic pain (CP), particularly with respect to the integral role of brain processes ^1^. While many studies have now thoroughly documented the effects of various CP conditions on both brain structure and function ^2,3^, much of this research has concentrated on gray matter alterations. Studies investigating structural changes to white matter (WM) in relation to chronic pain (CP) remain scarce and inconclusive ^4–8^. This is significant as white matter comprises nearly half of the brain ^9^, has importance in development ^10–12^, in function ^13–15^, in learning^16^ and is known to be impaired in numerous neurological conditions — including Alzheimer’s, Parkinson’s, depression, multiple sclerosis, and traumatic brain injury ^17–22^.

In line with trends in other scientific disciplines, the search for accurate biomarkers is a major focus within the pain research community. The increasing quality and availability of neuroimaging data makes it one of the most promising avenues for the development of biomarkers ^3,23–27^. Notably, longitudinal and machine learning approaches have yielded significant insights into brain characteristics that predict placebo response ^28^, longitudinal pain symptom change ^29^ and transition from subacute phase to CP ^30^.

Although the primary focus here is on the white matter, findings from other modalities can contextualize the study of WM in CP. Utilizing structural Magnetic Resonance Imaging (MRI) techniques, such as T1- and T2-weighted imaging, researchers have identified common and distinct brain features across various pain conditions ^31^. Even though recent meta-analysis report subtle, spatially distributed alterations in gray matter regions, including the amygdala, thalamus, hippocampus, insula, anterior cingulate cortex, and inferior frontal gyrus across CP conditions ^32,33^, unique “brain signatures” specific to individual pain conditions have also been observed. For example, patterns of gray matter density co-variation enabled the classification of individual brains to their condition – either chronic back pain (CBP), complex regional pain syndrome (CRPS) or knee osteoarthritis (OA) ^32^. Interestingly, these structural characteristics extend beyond the traditionally expected somatosensory regions, they are non-randomly distributed and may play a role to both the onset and maintenance of CP -- as seen in individuals transitioning from subacute to chronic low back pain (CLBP) who present smaller amygdala and hippocampi volumes ^34^. Collectively, these observations reinforce the utility of a nuanced and more global, condition-specific approach to chronic pain.

Studies examining functional MRI (fMRI), such as BOLD-weighted images, also indicate that certain dynamic features of brain activity, at rest or during task, are characteristic of various CP conditions. For instance, fibromyalgia patients show hypersensitivity to visual and pressure stimuli ^35–37^, whereas chronic lower back pain (CLBP) patients display distinct patterns of nucleus accumbens (NAc) activity in response to noxious stimuli ^38^. At the level of resting-state networks, fMRI studies have primarily emphasized changes in the default mode network, driven largely by sustained pain signaling via the medial prefrontal cortex^39^. Emerging research on dynamic connectivity further underscores the significance of looking at brain dynamics, revealing that dynamic features are more predictive of pain experiences than their static counterparts ^40^. Given these observations, examining the way white matter (WM) architecture constrains gray matter and functional alterations could offer valuable insights for the development and individualization of novel therapeutic interventions. Interestingly, some of these brain features appear to be plastic, displaying the capacity to partially reverse and reorganize either upon receiving treatment or as the condition progresses ^34,41,42^.

Diffusion magnetic resonance imaging (dMRI), which measures the signal loss due to the diffusion of water molecules in biological tissues as diffusion-sensitizing gradients are applied ^43^, stands as the most promising non-invasive tool for investigating structural changes in white matter. Introduced in the late 1980s, advancements in dMRI methodologies have been substantial ^44^, leading to increasingly sophisticated models for measuring white matter microstructure and reconstructing white matter connectivity. Early implementations primarily relied on calculating the Apparent Diffusion Coefficient (ADC), a scalar measure computed by averaging apparent diffusion on 1 to 3 directional dMRI scans ^45^. While these rapid scan sequences have become standard in clinical evaluations of specific pathologies like strokes and tumors, they lack specificity to define the microstructure and are rarely used for the evaluation of chronic pain patients. By the late 1990s, Diffusion Tensor Imaging (DTI) ^46^ was introduced, building on the limitations of ADC. By acquiring dMRI images in at least 6 directions, DTI can reconstruct diffusion directional preference, enabling the computation of metrics such as Fractional Anisotropy (FA) and Mean Diffusivity (MD). Nevertheless, in the context of tractography, DTI has limitations in capturing complex fiber configurations such as crossing or kissing fibers. To address this, High Angular Resolution Diffusion Imaging (HARDI) was developed in the early 2000s ^47^. By acquiring dMRI in at least 45 directions, HARDI was designed to resolve complex intravoxel structures.

Despite these advancements, the field of chronic pain (CP) has yet to yield the full potential of diffusion MRI-based methods. Diffusion tensor imaging (DTI) remains the most prevalent method in the field, so much so that it is frequently used as a synonym for dMRI in literature. However, for the community to update its methods, a cautionary note is warranted: if HARDI and multi-shell protocols gain widespread acceptance without standardization, there is a risk of exacerbating variability and noise in the CP literature. Therefore, the aim of this review is to critically assess the existing research concerning the characteristics of brain white matter revealed through diffusion MRI techniques. We examine impairments in white matter across various chronic pain conditions, as defined by the International Association for the Study of Pain (IASP), drawing upon findings from the 58 articles that met our inclusion criteria ^48,49^.

## Methods

### Information source

PubMed and Scopus databases were interrogated for articles dated up to 10^th^ March 2022. Articles were also searched using the Medical Subject Headings (MeSH) term on PubMed.

### Review strategy

This review follows the recommendations from the Preferred Reporting Items for Systematic reviews and Meta-Analyses extension for Scoping Reviews (PRISMA-ScR) Checklist. Search strategies were developed with a librarian of the Health Sciences Library of the Université de Sherbrooke. The keywords chosen for the review were: “magnetic resonance imaging”, “diffusion”, “pain”, and “brain”. The full search strategy can be viewed in the *supplementary material* (supp 1.).

### Study strategy and inclusion/exclusion criteria

The process for selecting studies in this investigation is depicted in Figure 1. First, from a collection of 447 papers, we discarded reviews and any articles that did not explicitly mention “diffusion,” “brain,” or “pain” in their title or abstract. Second, we eliminated case studies, non-English articles, and those in which pain was not the main focus. Third, we removed studies if they did not employ diffusion MRI (dMRI) to examine brain regions in individuals suffering from chronic pain or if the study population included participants under the age of one, veterans, amputees, recipients of deep brain stimulation (DBS), individuals with traumatic brain injury (TBI), postoperative patients, stroke or neurodegenerative disease-induced chronic pain sufferers, or subjects experiencing experimentally induced acute pain. Methodological articles and animal studies were also excluded. Fourth, we separated the remaining articles into three categories: chronic primary pain, chronic secondary pain based on the IASP classification ^48,49^, or articles missing information about the chronic pain condition, and only included articles on chronic primary pain. We further excluded studies that limited dMRI analysis solely to peripheral nerves, those that included only healthy control subjects, or those that failed to report results (a note on acute pain was also added in *supplementary material [supp 2.]*). Finally, an additional article was incorporated post hoc; this article had been referenced multiple times in the selected literature and was identified as pertinent due to its focus on the use of dMRI in chronic pain, despite not being retrieved by our initial search terms in its title or abstract.

**Figure 1.**
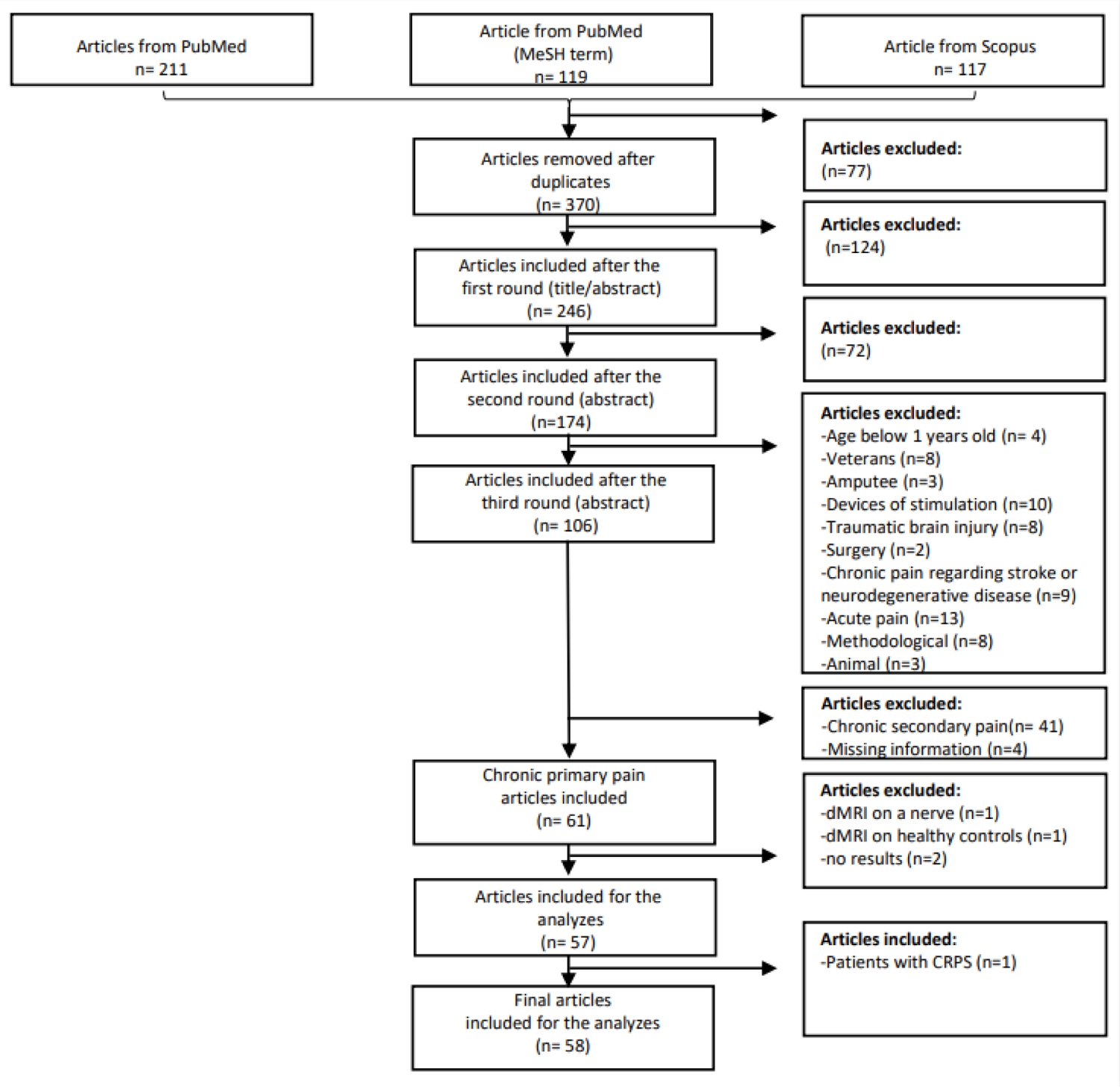
Flowchart of the article selection process, 58 final articles were included for the analyzes in this review.

### Study selection and analyses

The first three steps described in the section “*study strategy and inclusion and exclusion criterias*” were conducted by MS and PT; any disagreement was resolved between MS and PT. Subsequently, the classification into primary chronic pain and secondary chronic pain was made by MS and supported by PT and GLe. The remaining articles were then separated into five categories based on the IASP classification: (i) chronic widespread pain; (ii) complex regional pain syndrome; (iii) chronic primary headache or orofacial pain; (iv) chronic primary visceral pain and (v) chronic primary musculoskeletal pain. If applicable, half of the articles in each category were separated and respectively analyzed by MS and PT. The remaining articles were analyzed by MM and GLe. Finally, each reviewer (MS, PT, MM and GLe) extracted, based on the chart developed by PT, general study design information and dMRI-specific study data.

## Results

Of the 370 unique articles initially identified, 246 articles remained after removing reviews and articles which did not fit our inclusion criteria for the title and abstract. Subsequent removal of case-reports, articles not available in English and articles that only reported pain anecdotally yielded a total of 174 articles. Further exclusion of articles that did not acquire dMRI in brain regions or on a specified chronic pain condition population, left 106 articles. Then, the remaining articles were separated into three categories: chronic primary pain (61 articles), chronic secondary pain (41 articles) and articles missing information about the chronic pain condition (4 articles); only chronic primary pain articles were kept. Afterwards, we excluded articles where the dMRI was acquired: on a nerve, only on healthy participants and articles without results, leaving 57 articles. Finally, one previously omitted article was added retrospectively because it was cited several times by the remaining articles, giving a total of 58 articles for the final analysis. Figure 2 presents an interactive overview of 58 articles that met our inclusion criteria regrouped according to the latest IASP chronic pain definition and their analysis method.

**Figure 2.**
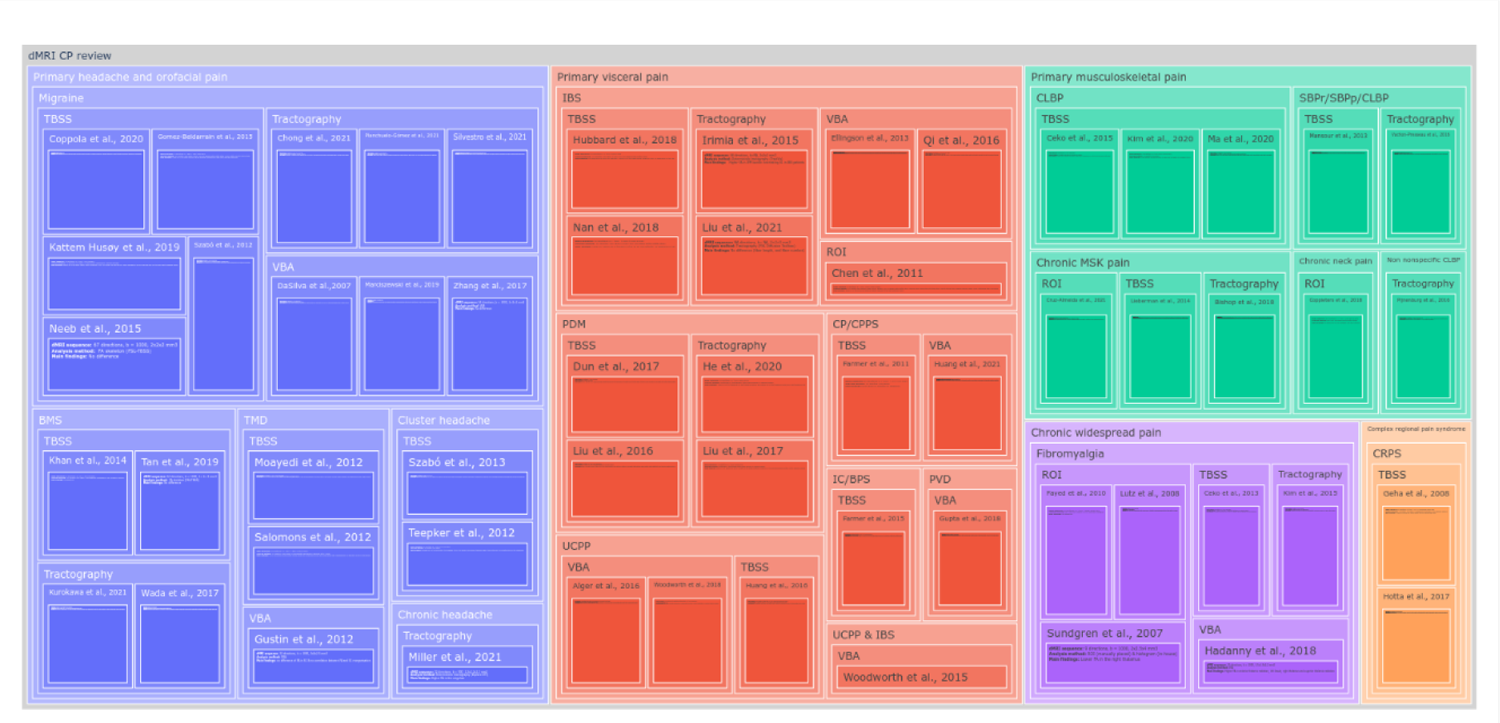
An interactive overview of 58 articles that met our inclusion criteria regrouped according to the latest IASP chronic pain definition and their analysis method. In **blue**: primary headache and orofacial, **red**: primary visceral pain, **green**: primary musculoskeletal pain, **purple**: chronic widespread pain and **orange**: complex regional pain syndrome. Dynamic figure can be found here: https://osf.io/4wyqt/

In the interest of clarity and conciseness, a list of all abbreviations used in the results is provided below (Table 2). Furthermore, abbreviations are redefined at the beginning of every section as not all chronic pain types may pertain to the interest of the reader.

**Table 1.**
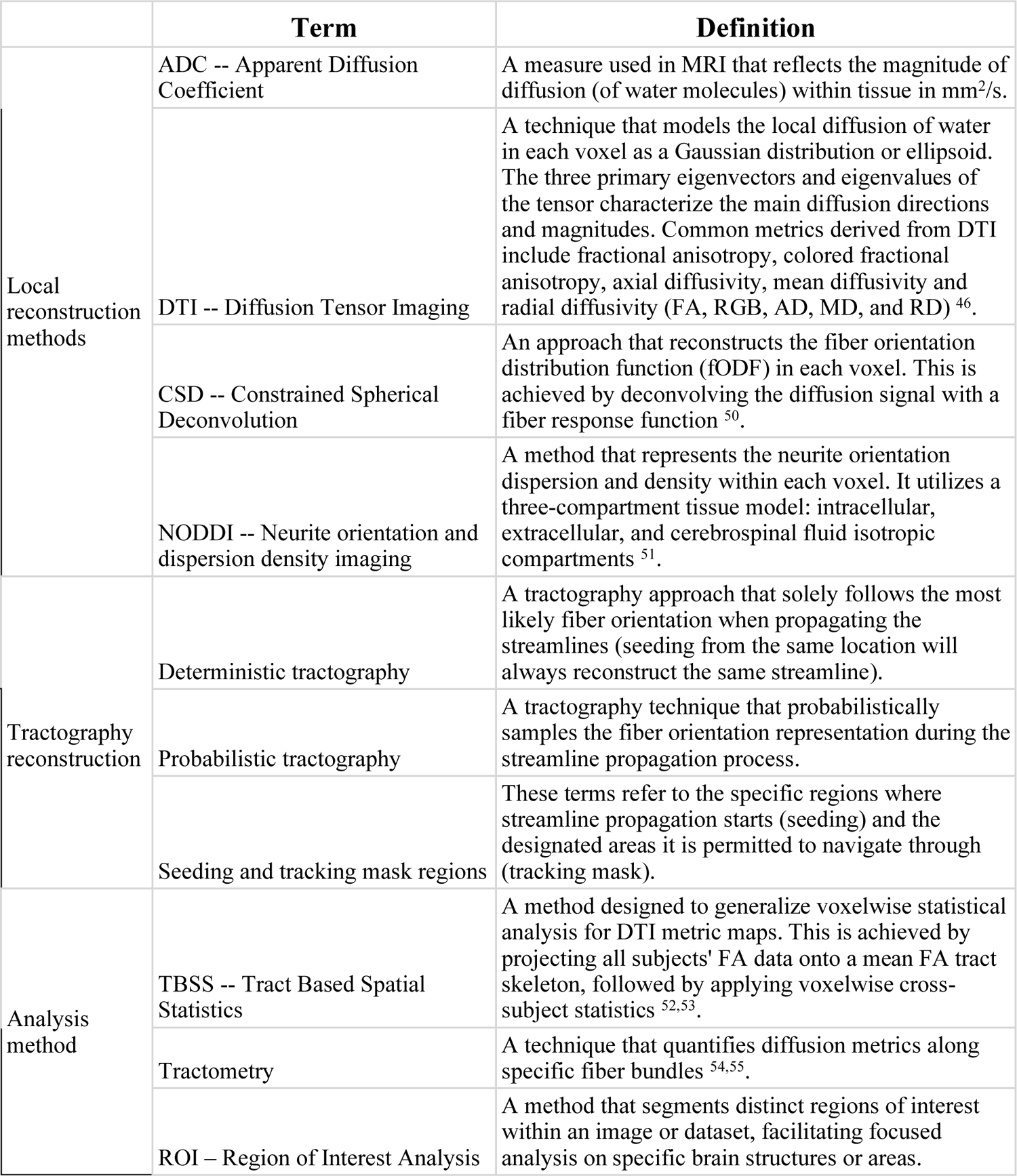
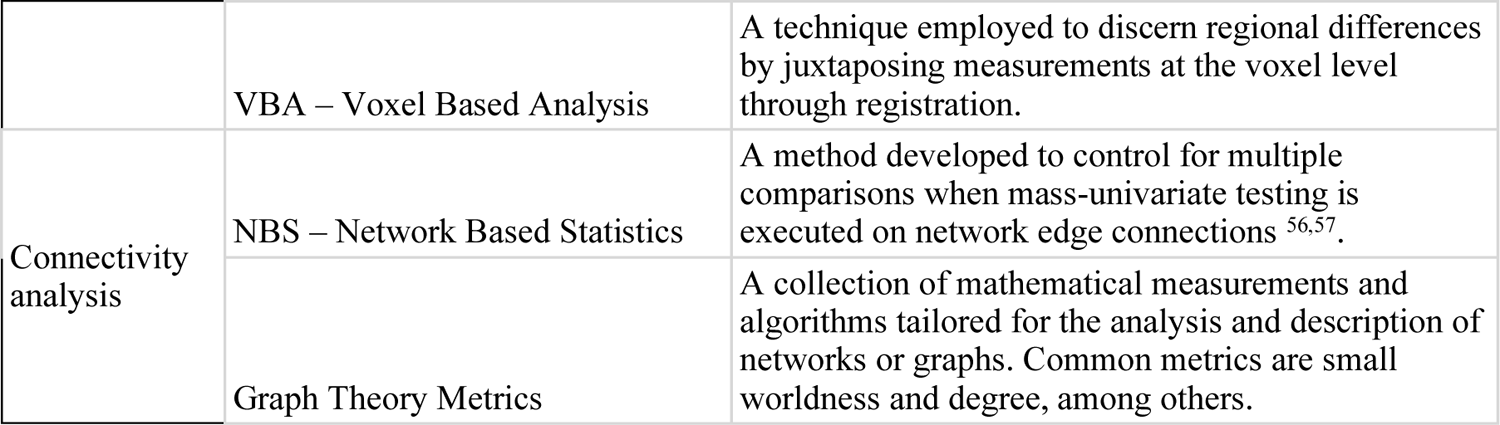
Glossary of technical dMRI terms.

|  | Term | Definition |
| --- | --- | --- |
| Local reconstruction methods | ADC -- Apparent Diffusion Coefficient | A measure used in MRI that reflects the magnitude of diffusion (of water molecules) within tissue in mm<sup>2</sup>/s. |
|  | DTI -- Diffusion Tensor Imaging | A technique that models the local diffusion of water in each voxel as a Gaussian distribution or ellipsoid. The three primary eigenvectors and eigenvalues of the tensor characterize the main diffusion directions and magnitudes. Common metrics derived from DTI include fractional anisotropy, colored fractional anisotropy, axial diffusivity, mean diffusivity and radial diffusivity (FA, RGB, AD, MD, and RD) <sup>46</sup>. |
|  | CSD -- Constrained Spherical Deconvolution | An approach that reconstructs the fiber orientation distribution function (fODF) in each voxel. This is achieved by deconvolving the diffusion signal with a fiber response function <sup>50</sup>. |
|  | NODDI -- Neurite orientation and dispersion density imaging | A method that represents the neurite orientation dispersion and density within each voxel. It utilizes a three-compartment tissue model: intracellular, extracellular, and cerebrospinal fluid isotropic compartments <sup>51</sup>. |
| Tractography reconstruction | Deterministic tractography | A tractography approach that solely follows the most likely fiber orientation when propagating the streamlines (seeding from the same location will always reconstruct the same streamline). |
|  | Probabilistic tractography | A tractography technique that probabilistically samples the fiber orientation representation during the streamline propagation process. |
|  | Seeding and tracking mask regions | These terms refer to the specific regions where streamline propagation starts (seeding) and the designated areas it is permitted to navigate through (tracking mask). |
| Analysis method | TBSS -- Tract Based Spatial Statistics | A method designed to generalize voxelwise statistical analysis for DTI metric maps. This is achieved by projecting all subjects' FA data onto a mean FA tract skeleton, followed by applying voxelwise cross-subject statistics <sup>52,53</sup>. |
|  | Tractometry | A technique that quantifies diffusion metrics along specific fiber bundles <sup>54,55</sup>. |
|  | ROI – Region of Interest Analysis | A method that segments distinct regions of interest within an image or dataset, facilitating focused analysis on specific brain structures or areas. |
|  | VBA – Voxel Based Analysis | A technique employed to discern regional differences by juxtaposing measurements at the voxel level through registration. |
| Connectivity analysis | NBS – Network Based Statistics | A method developed to control for multiple comparisons when mass-univariate testing is executed on network edge connections <sup>56,57</sup>. |
|  | Graph Theory Metrics | A collection of mathematical measurements and algorithms tailored for the analysis and description of networks or graphs. Common metrics are small worldness and degree, among others. |

**Table 2.**
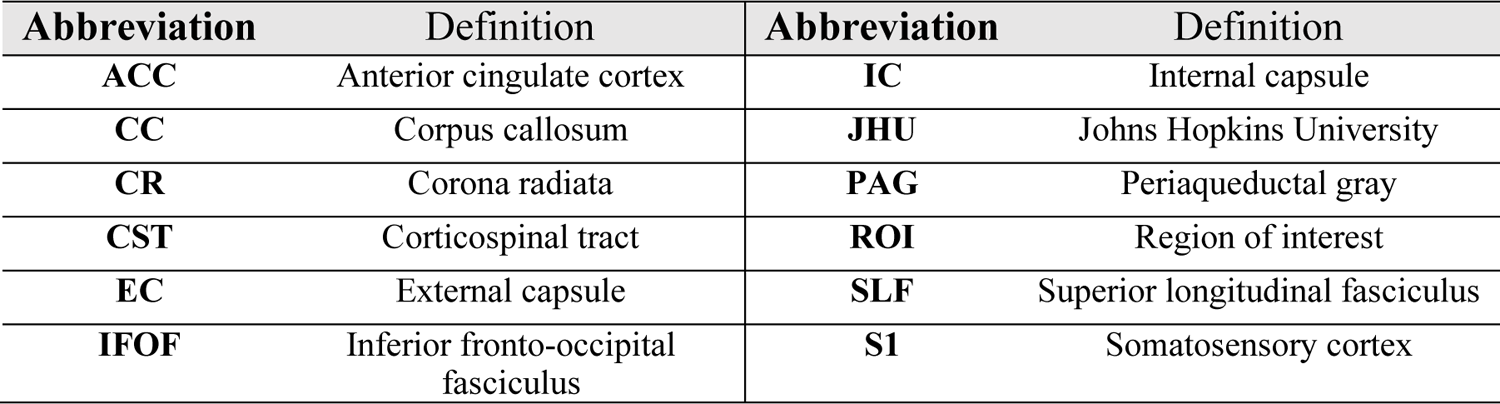
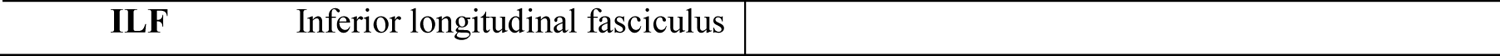
Abbreviations of regions of interest.

### Chronic primary headache or orofacial pain

The IASP classification defines chronic primary headache or orofacial pain as headache or orofacial pain manifesting for at least 15 days per month and persisting for over three months. The duration of untreated daily pain is a minimum of two hours or may manifest in multiple shorter episodes ^48^. In reviewing this category, 21 articles were identified: 11 focused on migraine ^58–68^, two on cluster headache ^69,70^, one on chronic headache ^71^, three on temporomandibular disorder (TMD) ^72–74^ and four on burning mouth syndrome ^75–78^.

In the studies examining **migraine**, five investigations employed FSL-TBSS for voxel-wise statistical analyses on whole-brain fractional anisotropy (FA) skeletons ^58,64–67^. While Neeb et al. ^64^ and Coppola et al. ^65^ solely relied on TBSS, Kattem Husøy et al. ^58^ and Szabó et al. ^67^ incorporated additional tractography analyses. Gomez-Beldarrain et al. ^66^ further extended their approach to include a region-of-interest (ROI) analysis, utilizing both white and gray matter atlases. Neeb et al. reported no significant differences in FA, mean diffusivity (MD), axial diffusivity (AD), and radial diffusivity (RD) between migraine patients and healthy controls. Coppola et al. examined both episodic and chronic migraines, finding no FA differences in episodic migraine patients but identifying higher RD and MD in bilateral superior and posterior corona radiata (CR), the bilateral genu of the corpus callosum (CC), the bilateral posterior limb of the internal capsule (IC) bilateral superior longitudinal fasciculus (SLF) in chronic migraine patients. Kattem Husøy et al. discovered elevated AD in major tracts, most notably in the bilateral CC, corticospinal tract (CST), inferior fronto-occipital fasciculus (IFOF), inferior longitudinal fasciculus (ILF), and left SLF. They also noted lower volumes in the CC and IFOF when employing deterministic tractography, but only in the new-onset headache group. Szabó et al. found lower FA and higher MD and RD values in frontal white matter bundles. Probabilistic tractography, originating from TBSS seed regions, revealed these bundles to be connected to the orbitofrontal cortex, insula, thalamus, and dorsal midbrain. Gomez-Beldarrain et al. observed lower FA in the TBSS skeleton and identified significantly different TBSS clusters between controls and migraine patients, which were mapped to ROIs in the Johns Hopkins University (JHU) diffusion tensor imaging (DTI)-based Atlas and the MNI structural Atlas. The interior insula, bilateral cingulate gyri, and right uncinate fasciculus were pinpointed as regions with lower FA values.

Three investigations exclusively utilized tractography ^60,63,68^. Chong et al. ^60^ employed probabilistic tractography to segment and average white matter microstructural properties with the aim of differentiating migraine patients from those with post-traumatic headaches. Their predictive model achieved an accuracy rate of 78%; however, no diffusion metrics were compared. Planchuelo-Gomez et al. ^63^ used probabilistic tractography to build a whole brain connectome by measuring streamline counts between gray matter regions. They observed both higher and lower numbers of streamlines in connections involving specific regions such as the superior frontal gyrus. Additionally, they found variations in FA, AD, and RD in connections involving regions like the hippocampus. Silvestro et al. ^68^ took a similar approach, employing probabilistic tractography to construct a whole-brain connectome. They quantified the connection probability between gray matter regions and further examined the resultant connectome using network-based statistics (NBS) and graph theory network analysis. Their findings highlighted nodes with significantly higher connection probabilities in multiple regions, including the precuneus, cuneus, amygdala, calcarine cortex, and posterior cingulate cortex, anterior cingulate cortex (ACC), postcentral gyrus, lingual and fusiform gyri, middle frontal gyrus and inferior and superior parietal lobules.

Three investigations utilized voxel-based analysis (VBA) ^59,61,62^. Zhang et al. ^59^ reported no significant differences using a whole-brain VBA approach via SPM12. On the other hand, Marciszewski et al. ^61^ observed elevated MD in various brain regions, including the spinal trigeminal nucleus, dorsal medial/lateral pons, midbrain periaqueductal gray (PAG), and cuneiform nucleus. They also identified higher FA in the white matter regions of the medial lemniscus and ventral trigeminal thalamic tract. Dasilva et al. ^62^ further extended their approach to include a ROI analysis, utilizing both white and gray matter atlases. They discovered lower FA values in specific patient subgroups: the ventro-lateral PAG was affected in migraine patients without aura, while the ventral trigemino-thalamic tract showed reduced FA in migraine patients with aura.

In the studies examining **cluster headache**, two investigations employed FSL-TBSS for voxel-wise statistical analyses on whole-brain FA skeletons ^69,70^. Szabo et al. ^69^ reported a significant increase in MD, AD, and RD across widespread white matter regions in the frontal, parietal, temporal, and occipital lobes. They also found reduced FA in the CC and in specific frontal and parietal white matter tracts, such as the bilateral forceps minor and major, the bilateral CR, the IC and external capsule (EC), the cerebral peduncle, the parietal juxtacortical white matter and the IFOF, predominantly on the contralateral side of the pain. Notably, AD exhibited a negative correlation with the frequency of headache attacks. Teepker et al. ^70^ reported alterations in FA in multiple brain regions, including the brainstem, thalamus, IC, superior and inferior temporal regions, frontal lobe, occipital lobe, and cerebellum.

In one study examining **chronic headache**, conducted by Miller et al. ^71^, they used deterministic tractography to segment and average diffusion metrics on tracts. They observed higher FA in the cingulum.

In the studies examining **temporomandibular disorders (TMD)**, two investigations employed FSL-TBSS for voxel-wise statistical analyses on whole-brain FA skeletons ^72,73^. Both studies extended their analysis to include tractography. Moayedi et al. ^73^ went further by also incorporating VBA. Salomons et al. ^72^ conducted their analysis using TBSS and then segmented major tracts via probabilistic tractography. They observed that FA values in connected white matter tracts along the CST were associated with feelings of helplessness; however, no group comparison of diffusion metrics were reported. Moayedi et al., after performing TBSS analyses, employed probabilistic tractography, using seed regions identified from significant TBSS clusters, to measure connection probability. They also applied a whole-brain VBA. Their findings indicated lower FA in the anterior limb of the IC and the EC. Additionally, they observed a higher connection probability from the CC to the frontal pole and a lower connection probability from the CC to the dorsolateral prefrontal cortex.

One investigation exclusively utilized whole-brain VBA ^74^. Gustin et al. ^74^ observed no significant differences in FA within the primary somatosensory cortex (S1), nor did they find any correlation between FA and reorganization of S1.

In the studies examining **burning mouth syndrome (BMS)**, two investigations employed FSL-TBSS for voxel-wise statistical analyses on whole-brain FA skeletons ^76,77^. Khan et al. ^76^ extended their study by incorporating probabilistic tractography to segment major white matter tracts. They found no significant differences in either TBSS-based diffusion metrics or average tract metrics and volumes. Tan et al. ^77^, who solely employed TBSS, also found no significant differences in the whole-brain FA skeleton.

Two investigations exclusively utilized tractography to construct whole brain connectomes ^75,78^. Both used probabilistic tractography to measure the streamline count between gray matter regions and subsequently applied graph theory network analysis methods to further examine the connectomes. Wada et al. ^75^ observed localized changes in connectivity within the ACC and prefrontal cortex, specifically in the medial orbitofrontal cortex and pars orbitalis. They also noted strengthened connections between the ACC and medial prefrontal cortex with regions such as the basal ganglia, thalamus, and brainstem. Despite these findings, no significant differences were observed in graph theory metrics. Kurokawa et al. ^78^ reported increased connectivity and betweenness centrality in the left insula, right amygdala, and right lateral orbitofrontal cortex as well as reduced betweenness centrality and connectivity in the right inferotemporal cortex.

### Chronic primary visceral pain

The IASP classification defines chronic primary visceral pain as persistent or recurrent pain (for longer than three months) that occurs in the internal organs of the head (or neck) region and of the thoracic, abdominal, or pelvic cavities, unexplained by any other condition ^48^. The pain anatomical location corresponds with typical referral pain patterns from specific internal organs. It can manifest in diverse forms, such as pain in the digestive system, the thoracic region, and the abdominal region, as well as pelvic pain originating from the viscera of the digestive, urinary, and genital systems. In this category, 19 studies were included. Among them, four focused on primary dysmenorrhea (PDM) ^79–82^, seven on irritable bowel syndrome (IBS) ^83–89^, four on urologic chronic pelvic pain (UCPP) ^90–93^, two on prostatitis/chronic pelvic pain syndrome (CCP/CPPS) ^94,95^, one on cystitis/bladder pain syndrome ^96^ and one on provoked vestibulodynia (PVD) ^97^.

In the studies examining **primary dysmenorrhea (PDM)**, two investigations employed FSL-TBSS for voxel-wise statistical analyses on whole-brain fractional anisotropy (FA) skeletons ^79,82^. While both studies utilized TBSS, Liu et al. ^82^ extended their analysis to include tractography. Dun et al. ^79^ observed lower FA values, alongside higher mean diffusivity (MD) and radial diffusivity (RD), across various white matter fibers tracts. These tracts included the splenium part of the corpus callosum (CC), the posterior limb of the internal capsule (IC), the superior and posterior of the corona radiata (CR), as well as the posterior thalamic radiation. Conversely, Liu et al. reported higher FA and lower MD and RD values, primarily in the CC, fornix, bilateral IC, bilateral external capsule (EC), CR, and bilateral posterior thalamic radiation, bilateral sagittal stratum, right cingulum and bilateral superior longitudinal fasciculus (SLF). In addition, tractography was employed by Liu et al. to visually inspect connectivity originating from TBSS seed regions.

Two investigations exclusively utilized tractography ^80,81^. Both studies performed tractography in a population atlas to manually segment white matter tracts of interest. Subsequently, these segmentations were registered to the native space for a region-of-interest (ROI) analysis of diffusion tensor imaging (DTI) metrics. He et al. ^80^ reported lower FA values in connections between the thalamus and somatosensory cortex (S1) as well as between the thalamus and insula. Conversely, they found higher FA values in the tracts connecting the thalamus to the dorsal anterior cingulate cortex (ACC) and the supplementary motor area. Liu et al. ^81^ observed lower FA and axial diffusivity (AD), along with higher RD and MD, in a specific cluster located in the dorsal posterior cingulum and in the parahippocampal segment of the cingulum bundle.

In the studies examining **irritable bowel syndrome (IBS)**, three investigations employed FSL-TBSS for voxel-wise statistical analyses on whole-brain FA skeletons ^86–88^. Chen et al. ^87^ further refined their TBSS analysis with a ROI approach using multiple atlases, including the Johns Hopkins University (JHU) white matter atlas, Harvard-Oxford cortical structural atlas, MNI structural atlas, Talairach Daemon labels, and the Juelich histological atlas. Similarly, both Hubbard et al. ^86^ and Nan et al. ^88^ employed an ROI-based approach, albeit solely using the JHU white matter atlas for segmentation. Chen et al. observed higher FA values in the fornix and EC adjacent to the right posterior insula. Hubbard et al., while not finding any significant differences in the whole FA skeleton, did report lower FA in the lower dorsal cingulum; however, they found no variations in MD and RD values. Nan et al. reported lower FA and higher RD specifically in the genu of the CC, with no observed differences in MD.

Two studies exclusively employed tractography methods ^83,89^. Irimia et al. ^83^ utilized deterministic tractography to calculate average tract DTI metrics while Liu et al. ^89^ applied tractography originating from the bilateral posterior cingulate gyrus to compute metrics such as average tract FA, fiber length, and streamline count. Irimia et al. observed higher FA values in white matter bundles innervating the S1. In contrast, Liu et al. did not report any significant differences in their measured parameters.

Two studies employed voxel-based analysis (VBA) ^84,85^. Ellingson et al. ^84^ extended their analysis by incorporating a ROI approach using the JHU atlas and applied probabilistic tractography to compute the number of streamlines between certain atlas ROIs. Qi et al. ^85^ similarly extended their analysis by applying tractography but originated it from fMRI-defined ROI clusters to compute average tract DTI metrics, streamline count, and path length. Ellingson et al. observed higher FA values in various grey matter regions including the globus pallidus, putamen, medial thalamus, and sensory and motor cortices as well as in white matter regions such as the primary cortical projections from the thalamus, posterior cingulate, frontal lobe and ACC white matter and CC when using VBA. They also reported higher MD within the globus pallidus, IC, thalamus, CR, and areas connected to sensory, pre-frontal, and posterior parietal regions. In terms of tract density, they found higher values in tracts connecting the thalamus to the prefrontal cortical regions and the medial dorsal nuclei to the ACC. Conversely, they observed lower tract density in connections between the globus pallidus and the thalamus. In contrast, Qi et al. did not find any significant differences in path length, tract count, or FA within the fibers connecting the bilateral ventral ACC to the inferior parietal lobules.

In the studies examining **urologic chronic pelvic pain (UCPP)**, one study conducted by Huang et al. ^91^ utilized FSL-TBSS for voxel-wise statistical analyses on whole-brain FA skeletons. Huang et al. extended their TBSS analysis by incorporating a ROI approach using the JHU white matter atlas. Huang et al. reported lower FA and higher AD values in the thalamic radiation but found no significant differences in either MD or RD.

Three studies employed VBA ^90,92,93^. The 2018 study by Woodworth ^92^ focused solely on whole-brain VBA, while the 2015 Woodworth study ^90^ extended its analysis to include probabilistic tractography. Alger et al. ^93^ incorporated a ROI approach using the JHU white matter atlas. In the 2018 study, Woodworth reported significant correlations between DTI measures and urinary protein quantifications; however, no comparative analyses were conducted between different groups in terms of diffusion metrics. The 2015 Woodworth study found lower FA, lower generalized FA, lower tract density, and higher MD in brain regions typically associated with the perception and integration of pain information. Interestingly, the study did not compare IBS patients to a control group but did find them to be significantly different from UCPP patients. Alger et al. primarily aimed to evaluate the variability in FA measurements. Their study obtained data from various acquisition sites for neurologic chronic pain syndrome in healthy controls but did not conduct any comparative analyses on diffusion metrics.

In the studies examining **prostatitis/chronic pelvic pain syndrome (CCP/CPPS)**, one study utilized FSL-TBSS for voxel-wise statistical analyses on whole-brain FA skeletons^94^. Farmer et al. ^94^ reported no significant differences in whole-brain FA skeletons across various DTI measures.

In a single study conducted by Huang et al. ^95^, both VBA and tractography were employed to calculate whole-brain connectome and graph theory metrics. Huang et al. observed lower global efficiency in the right middle frontal gyrus (orbital part) and higher global efficiency in the left middle cingulate and paracingulate gyrus. Additionally, they reported increased local efficiency in the left middle cingulate and paracingulate gyri, as well as the paracentral lobule.

In the study examining **cystisis/bladder pain syndrome**, a study utilized FSL-TBSS for voxel-wise statistical analyses on whole-brain FA skeletons ^96^. Farmer et al. ^96^ reported lower FA values in specific regions: the right thalamic radiation, the left forceps major, and the right longitudinal fasciculus. Conversely, they observed higher FA in the right SLF as well as in the bilateral inferior longitudinal fasciculus (ILF).

In the study examining **provoked vestibulodynia (PVD)**, a study conducted by Gupta et al. employed VBA on the whole brain and a ROI approach with the Harvard-Oxford subcortical atlas to segment gray matter regions ^97^. The investigators reported extensive increases in FA within the somatosensory and basal ganglia regions as well as variations in MD specifically in the basal ganglia.

### Chronic primary musculoskeletal pain

The IASP classification defines chronic primary musculoskeletal pain as persistent or recurrent (for longer than three months) pain that occurs in the muscles, bones, joints, or tendons ^48^. A classic example is chronic primary low-back pain. This category is further subclassified based on pain anatomical location, including the upper back as chronic primary cervical pain, the mid-back as chronic primary thoracic pain, the lower back as chronic primary low-back pain, and the limbs as chronic primary limb pain. In this category, ten studies were included. Among them, five focused on sub-acute/chronic low back pain (SBP/CLBP) ^34,98–101^, three on chronic musculoskeletal pain syndrome ^102–104^, one on nonspecific low back pain ^105^ and one on chronic neck pain ^106^.

In the studies examining **sub-acute/chronic low back pain (SBP/CLBP)**, four studies employed FSL-TBSS to conduct voxel-wise statistical analyses on whole-brain fractional anisotropy (FA) skeletons ^98–101^. Among these, Kim et al. ^101^ solely employed TBSS, Mansour et al. ^98^ extended their analysis to include tractography, and both Ma et al. ^99^ and Ceko et al. ^100^ applied a region of interest (ROI) approach. Specifically, Ma et al. utilized a white matter atlas for their ROI analysis, while Ceko et al. employed ROI clusters derived from a prior fMRI study. Kim et al. observed that CLBP patients exhibited reduced FA in both the somatosensory cortex (S1)-back and S1-finger regions when compared to controls by averaging FA skeleton diffusion tensor imaging (DTI) metrics in ROIs. Mansour et al. found lower FA values in three distinct clusters: one in the temporal part of the superior longitudinal fasciculus (SLF), a second in the left retro-lenticular part of the internal capsule (IC), and a third involving the left anterior limb of the IC as well as portions of the corpus callosum (CC), including the anterior corona radiata (CR) in SBP patients with pain that persisted compared to SBP patients that recovered after a year. Ma et al. reported lower FA in several regions: the CC, bilateral anterior and right posterior thalamic radiation, right SLF, and left anterior CR among CLBP patients. Contrarily, Ceko et al. didn’t find any whole-brain DTI metric differences in the FA skeleton, but instead measured an increased FA in the left insula post-treatment.

One study exclusively used probabilistic tractography to reconstruct the whole brain connectome and evaluate the number of connections and connection probabilities ^34^. Vachon-Presseau et al. ^34^ observed higher density of connections between corticolimbic regions, such as the nucleus accumbens, the amygdala, the hippocampus and the prefrontal cortex in patients with pain that persisted compared to SBP patients that recovered after a year. No comparisons were made with respect to DTI metrics.

In the studies examining **non-specific chronic low back pain (NSCLBP)**, Pijnenburg et al. ^105^ study exclusively used probabilistic tractography to reconstruct the whole brain connectome and evaluate graph theory network analysis. The investigators observed lower local efficiency in NSCLBP cases; notably, no comparative analysis was conducted on diffusion metrics.

In the studies examining **chronic musculoskeletal pain**, two studies employed FSL-TBSS to conduct voxel-wise statistical analyses on whole-brain FA skeletons ^102,103^. Lieberman et al. ^103^ extended their analysis by incorporating a ROI approach using a white matter atlas. Bishop et al. ^102^ also employed an ROI approach using the JHU white matter atlas, a whole brain fixel-based analysis and a probabilistic tractography with constrained spherical deconvolution (CSD) reconstruction of the whole-brain connectome to apply network-based statistics (NBS). Lieberman et al. found no significant differences in FA or axial diffusivity (AD) in the whole-brain TBSS FA skeleton. However, they reported higher radial diffusivity (RD) in multiple regions, including the body of the CC, right SLF and both anterior and posterior limbs of the IC. Further ROI analyses revealed lower FA in the splenium of the CC and the left temporal lobe branch of the cingulum bundle adjacent to the hippocampus. Elevated RD was also found in several regions, such as the splenium of the CC, the right limbs of the IC, part of the external capsule (EC) adjacent to the insular cortex, the SLF and the cerebral peduncle. Bishop et al. observed lower F1 values in the right EC, right SLF, and right uncinate fasciculus, as well as lower F2 values in the left cingulum and the splenium of the CC (F1 and F2 are respectively the first and second fiber population partial volume fractions, these diffusion properties can only be calculated when using advanced diffusion models that can account for multiple crossing fibers within a voxel). They also found lower mode of anisotropy in several areas including the splenium of the CC, left cerebral peduncle, and bilateral EC. In their whole-brain TBSS skeleton analysis, no differences in F1 and F2 were found. However, fixel-based analysis revealed reduced fiber density in the splenium of the CC and the right temporal lobe white matter region of the inferior frontal-occipital fasciculus (IFOF). No differences were observed in fiber cross-section. Utilizing a connectome approach, Bishop et al. reported increased connectivity between several regions, including the hippocampus, parietal cortex, thalamus, precuneus, and visual cortex structures like the calcarine and cuneus gray matter. One study exclusively used an ROI approach, utilizing DTI and neurite orientation dispersion and density imaging (NODDI) metrics ^104^. By targeting ROIs within both white matter and gray matter atlases, Cruz-Almeida et al. ^104^ observed lower orientation dispersion index values in the white matter of several regions, including the anterior CR, right posterior thalamic radiation, uncinate fasciculus, superior cerebellar penduncle, and fornix.

In a study focused on **chronic neck pain**, conducted by Coppieters et al. ^106^, an ROI approach using a white matter atlas was employed to assess DTI metrics. The investigation revealed no significant differences in ROI-based DTI metrics between patients with chronic neck pain and healthy controls.

### Chronic widespread pain

The IASP classification defines chronic widespread pain as persistent or recurrent (for longer than three months) diffuse musculoskeletal pain that occurs in a minimum of four body regions and in at least three of four body quadrants (upper–lower/left–right side of the body) ^48,107^. In this category, six studies were included. All of which focused on fibromyalgia (FM) ^108–113^.

In the studies focused on fibromyalgia, one study conducted by Ceko et al. ^109^ utilized FSL-TBSS for voxel-wise statistical analyses on whole-brain fractional anisotropy (FA) skeletons. Additionally, they employed tractography to examine connectivity originating from significant TBSS clusters. While no differences were observed in whole-brain FA skeletons with respect to diffusion tensor imaging (DTI) measures, the investigators did report lower FA in regions adjacent to areas where significant gray matter volume differences were identified.

One study exclusively used tractography to construct whole brain connectomes using streamline counts between gray matter regions ^110^. Kim et al. ^110^ used probabilistic tractography to build a whole brain connectome by measuring streamline count between gray matter regions. Notably, they found no differences in the white matter fiber count connecting areas associated with hyperalgesia and clinical pain (no local diffusion metrics were compared).

One study exclusively used a whole brain voxel-based analysis (VBA) ^111^. Hadanny et al. ^111^ observed higher FA in several regions, including the anterior thalamic radiation, left insula, right thalamus, and superior thalamic radiation.

Three studies exclusively used region of interest (ROI) based analysis methods ^108,112,113^. Sundgren et al. ^108^ used ROI masks for the whole brain and manually placed ROI spheres to utilize a histogram comparison method. Their findings included lower FA in the right thalamus, with no significant results for apparent diffusion coefficient (ADC). Fayed et al. ^112^ also manually positioned ROI spheres to compare FA and ADC metrics but found no significant differences between groups. Lutz et al. ^113^ manually segmented ROIs to compare FA and ADC metrics. They observed lower FA in both the thalami, thalamocortical tracts, and both insular regions, alongside higher FA in the postcentral gyri, amygdala, hippocampi, superior frontal gyri, and anterior cingulate gyri. No significant differences were found in ADC metrics.

### Complex regional pain syndrome (CRPS)

The IASP classification defines complex regional pain syndrome (CRPS) as persistent, or recurrent (for longer than three months) pain characterized by its regional distribution and time course. The pain typically begins distally in an extremity following trauma and is disproportionate in both magnitude and duration when compared to the usual course of pain after similar tissue injuries ^48,114,115^. While two subtypes of CRPS have been identified, they are beyond the scope of this review. In this category, two studies were included ^116,117^.

In the studies focused on **Complex regional pain syndrome (CRPS)**, two studies employed FSL-TBSS for voxel-wise statistical analyses on whole-brain fractional anisotropy (FA) skeletons ^116,117^. While both studies used TBSS, Geha et al. ^117^ extended their approach to include probabilistic tractography, which was applied to investigate connectivity originating from voxel-based morphometry (VBM) seed regions. Geha et al. reported a lower FA cluster within the left callosal fiber tract. Hotta et al. ^116^, who solely utilized TBSS, observed higher mean diffusivity (MD), axial diffusivity (AD), and radial diffusivity (RD) in the genu, body, and splenium of the corpus callosum (CC) as well as in the left anterior, posterior and right superior parts of the corona radiata (CR). Across the whole-brain FA skeletons, they observed average lower FA, higher MD and RD, and no differences were observed in AD.

### Summary of our findings

To illustrate the results covered in this review, we have provided a summary of the types of chronic pain, the analysis methods and the reported regions and tracts used for the study of chronic pain with diffusion MRI in figure 3.

**Figure 3.**
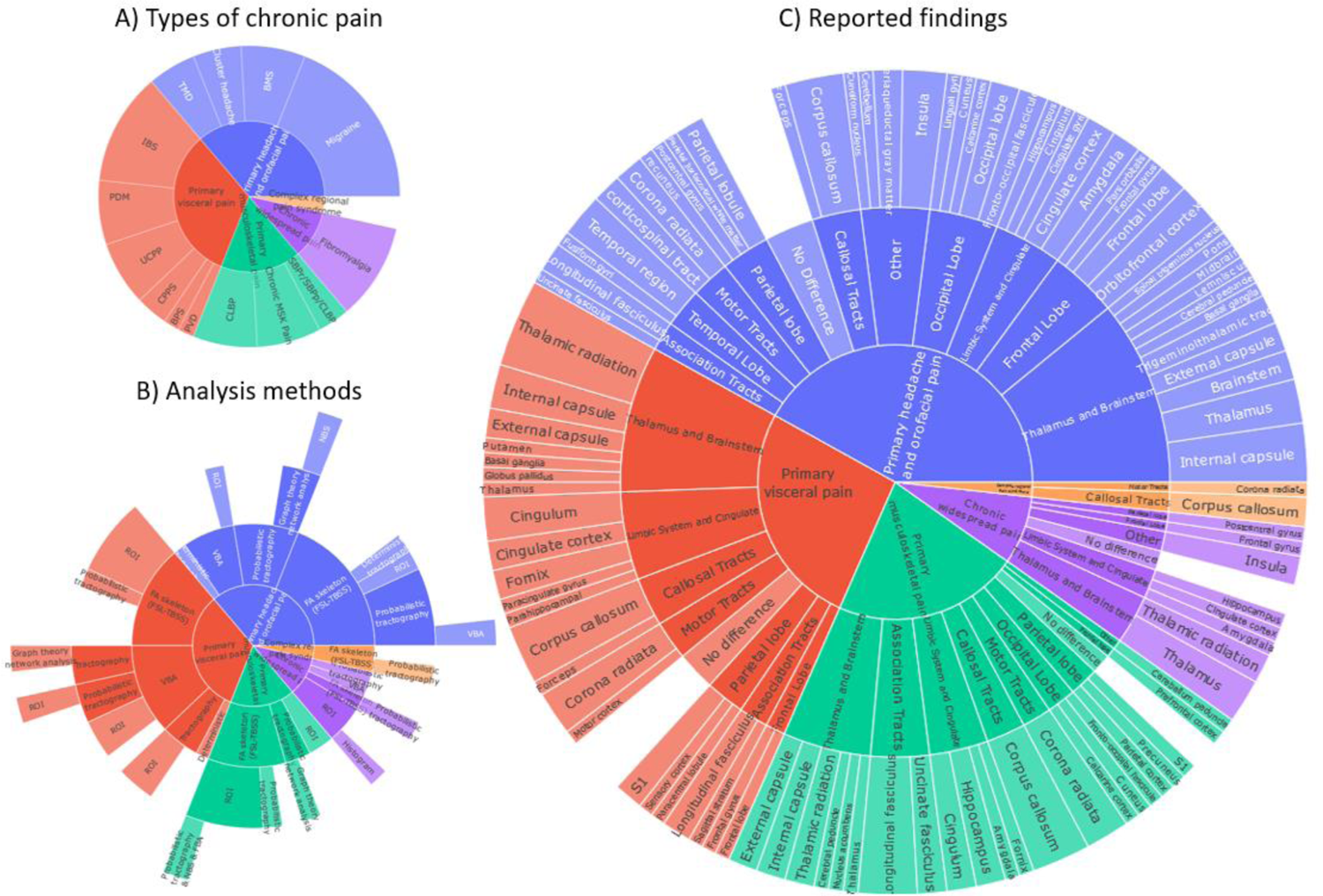
An interactive visual summary of the reviewed studies in terms of the types of chronic pain studied (A), the used analysis methods (B) and the tracts and regions reported as significant findings (C). In **blue**: primary headache and orofacial, **red**: primary visceral pain, **green**: primary musculoskeletal pain, **purple**: chronic widespread pain and **orange**: complex regional pain syndrome. Dynamic figure can be found here: https://osf.io/4wyqt/

## Discussion

The purpose of this review was to provide a critical summary of the use of brain dMRI for the study of primary chronic pain conditions. Each article was classified according to the latest IASP chronic pain definition, dMRI sequence and analysis method. The main findings of this review highlight the difficulty of delineating common white matter abnormalities for each chronic pain condition. Indeed, as shown by figure 4, sixty-four percent (35/55) of all reported regions/tracts are only reported once or twice across all studies. Furthermore, the variety of reported metrics for a given region accentuates the lack of consensus of white matter properties for each chronic pain condition. This observation comes in part from the vast number of possibilities to analyze and further report results from dMRI data. Notwithstanding, some regions are reported more consistently. For example, the corpus callosum is reported in twenty-four percent (14/58) of all studies and a few tracts and regions emanating from the thalamus are reported in over 10% of all studies. However, these findings must be interpreted with caution as these regions could either be relatively easier to investigate (due to size, shape and localization), more common in the pain literature and subject to “publication bias” ^118^ as the average study CP subjects sample size is relatively low (37 subject/study)^118^.

**Figure 4.**
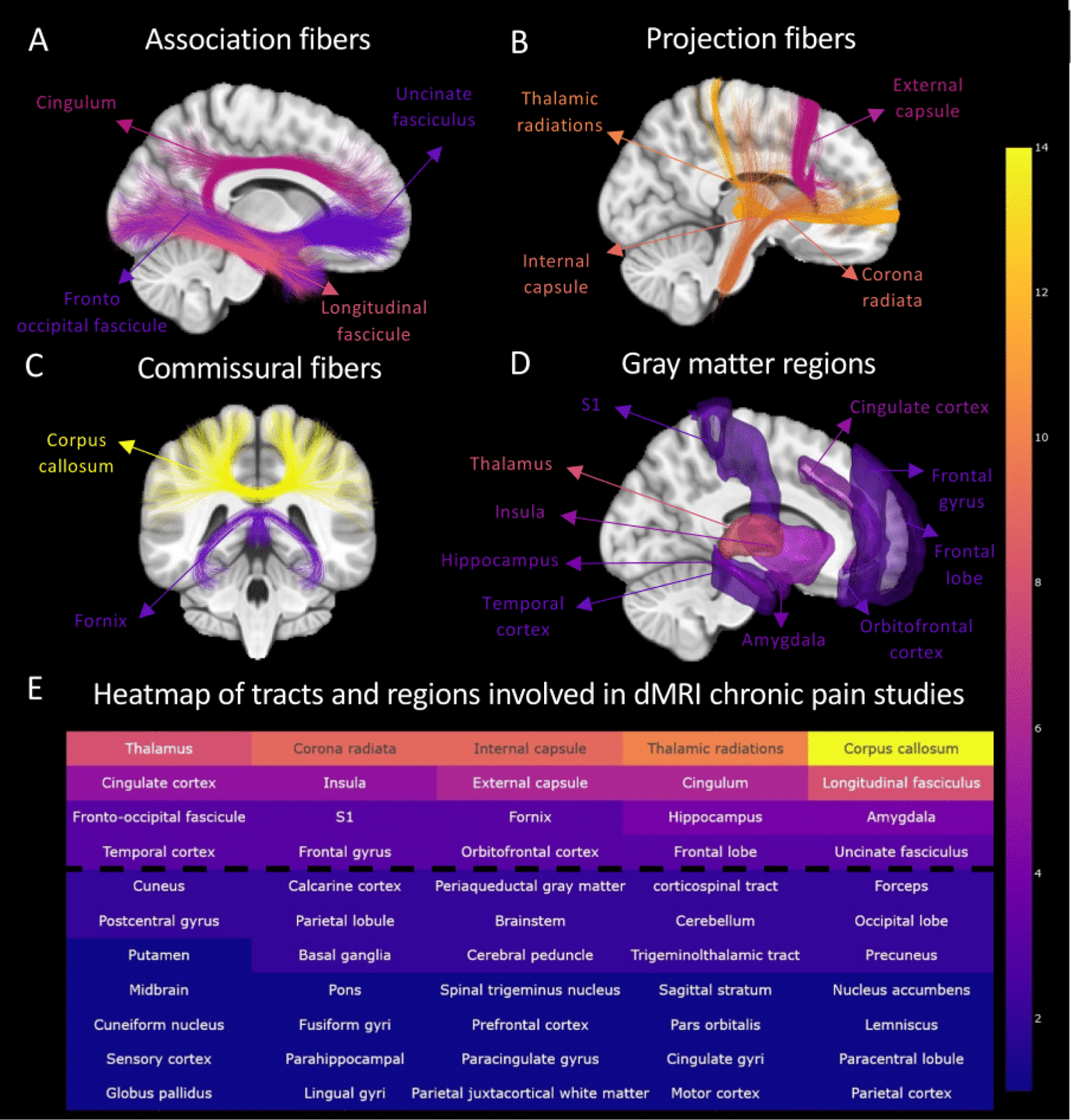
Visual representation of the association fibers (A), the projection fibers (B), the commissural fibers (C) and the gray matter regions (D) which were mentioned by at least three different articles in this review (above the dashed line in the heatmap). (E) An interactive heatmap of how consistently each tract and region is reported as significant findings across the studies of this review. The tracts and regions with warm colors (towards yellow) are reported more consistently (with a maximum of 14 studies that report the corpus callosum) relatively to tracts and regions with cold colors (towards blue; with over 50% of regions mentioned only once or twice). Dynamic heatmap can be found here: https://osf.io/4wyqt/

Seventy-nine percent of studies (46/58) have used DTI metrics group comparisons between CP and healthy controls as part of their reported findings. While this seems to be a common first step, thirty-one percent of studies (18/58) reported further analyses than DTI metric group comparisons (seven articles in the orofacial pain category, seven articles in the visceral pain category, three articles in the musculoskeletal pain category and one article in the widespread pain category). These additional analysis methods are highlighted in figure 5 to illustrate the diversity of dMRI analysis approaches.

**Figure 5.**
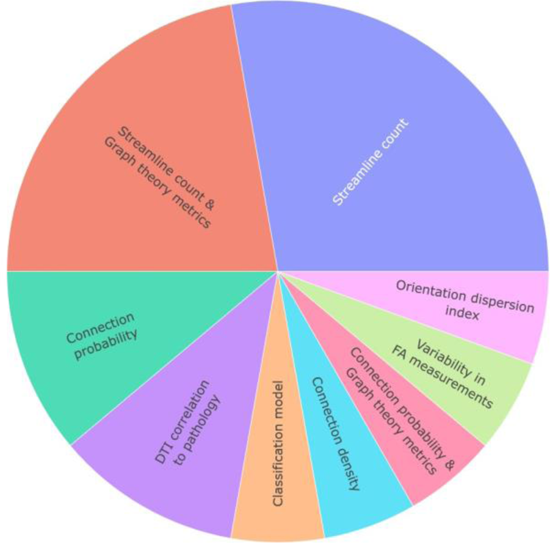
Distribution of alternative metrics to diffusion metrics comparisons between chronic pain (CP) and healthy controls used in diffusion MRI. Dynamic figure can be found here: https://osf.io/4wyqt/

**Figure 6.**
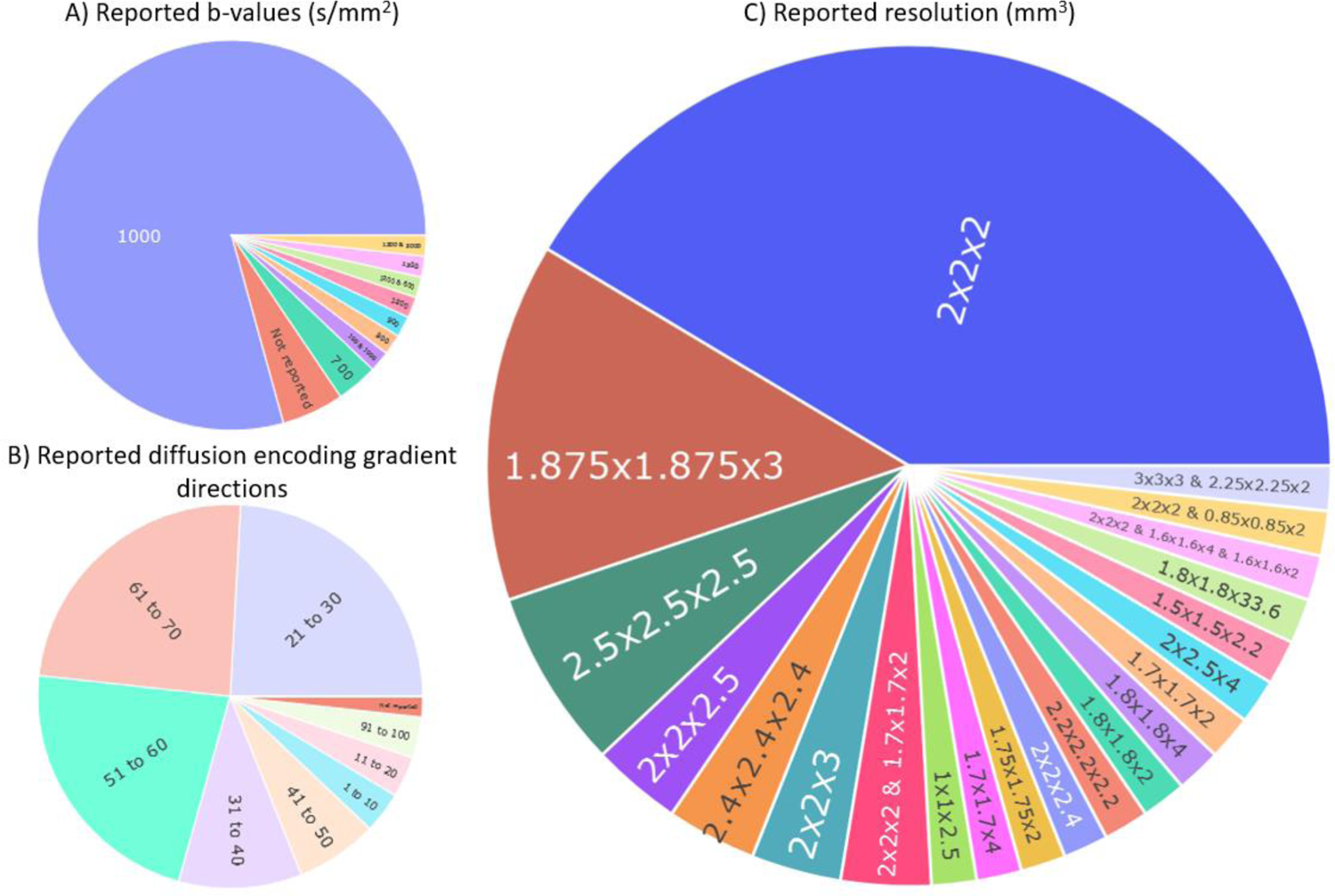
Overview of the different acquisition parameters used in the studies of the review. Distribution of the b-value in s/mm^2^ (A), the number of diffusion encoding gradient directions (B) and the resolution in mm^3^ (C). Dynamic figure can be found here: https://osf.io/4wyqt/

Interestingly, twenty-four percent of the studies (14/58) reported no differences between the groups that were investigated. Of these, five were in the orofacial pain category (two migraine, one TMD and two BMS studies), two in the musculoskeletal pain category (one CLBP and one chronic neck pain), five in the visceral pain category (three IBS, one UCPP and one CP/CPPS) and two on widespread pain (fibromyalgia).

Finally, six percent of the studies (4/59) reported differences of FA or ODI in the fornix (two in visceral pain, one in CRPS and one in MSK) even though, due to its unique location surrounded by CSF, it is most likely affected by partial volume effects (PVE) even when using state-of-the art dMRI acquisition sequences ^119^. Furthermore, as presented later, when using a TBSS approach, the fornix is almost absent from the TBSS FA skeleton.

## Main critics of the approaches used in the 58 articles

### Acquisition parameters

Diffusion MRI metrics, though quantitative, are somewhat restricted in terms of their sensitivity and specificity. This necessitates careful interpretation of these metrics, considering methodological, technical, and biological factors. A notable issue affecting the consistency and reliability of dMRI is the substantial variation in scanning parameters across different studies. For instance, a recent study explored the variability in diffusion-weighted MRI across multiple sites, scanners, and subjects. The study demonstrated that, under specific acquisition conditions, the variability between different scanners assessing the same subject could be comparable to the variability between different subjects assessed on the same scanner ^120^. As for any MRI sequences, many parameters need to be properly chosen to ensure high-quality diffusion-sensitive images. Three key parameters significantly influence both dMRI image quality and the outcomes of their subsequent analyses: 1) the b-value; representing the strength, duration and timing of the diffusion encoding gradients ^43,121^; 2) the number of diffusions encoding gradient directions; generally representing the number of diffusion gradient directions applied over a sphere ^122,123^; and 3) the voxel dimensions; representing the length, width and height of the 3D image voxels ^124^. For reference, the most common acquisition parameters used for DTI are in the range of a b-value of 1000 s/mm^2^, 30 unique gradient directions and a 2mm isotropic resolution.

1. Across studies, the b-value parameter was the most stable with ∼75% of studies using a b-value of 1000 s/mm^2^. However, some studies reported b-values in an unusual range— such as 700, 800, 900, 1200, and 1300 s/mm^2^—without providing any explanation. Additionally, certain studies failed to report the b-value at all. Strikingly, only two groups performed multi-shell acquisitions, an acquisition strategy introduced over 15 years ago ^125^, which allows for state-of-the-art dMRI analysis. Using modern multiband sequences, multi-shell images for whole brain in-vivo imaging can be acquired in an acceptable amount of time -- between 10 and 20 minutes at most.
2. When investigating the number of diffusion encoding gradient directions, we found that approximately 55% of the studies acquired a minimum of 30 directions (with a maximum of 99 directions). This parameter, however, exhibited significant variability; some studies used as few as six or nine directions. This is of concern because the theoretical minimum required to adequately describe a diffusion tensor is six gradient directions ^126,127^. In the broader context of tractography and fiber orientation estimation, high angular resolution diffusion imaging (HARDI) has emerged as an effective acquisition strategy designed to address the limitations of traditional diffusion tensor imaging (DTI) ^128,129^. Based on the acquisition of over 50 gradient directions at a single high b-value, HARDI (even when paired with a multi-shell acquisition) can be completed in a reasonable timeframe, between 10 and 20 minutes at most.
3. Lastly, when investigating image resolution, the overall voxel volume ranged from 2.4mm^3^ to 20mm^3^. While forty-seven percent of studies used the conventional 2mm isotropic resolution, there was notable variability, with some studies acquiring highly anisotropic voxels -- such as 1.875×1.873×3 or 2×2.5×4mm^3^. These anisotropic voxel dimensions can introduce biases into diffusion magnetic resonance imaging (dMRI) analyses. For instance, larger voxels are more likely to contain multiple fiber populations, thereby reducing fiber orientation homogeneity and affecting diffusion metrics ^124^. Additionally, anisotropic voxels can influence tractography algorithms, particularly in situations involving branching fibers^130^.

Overall, the observations made in this section highlight the heterogeneity of acquisition parameters and pre-processing pipelines across chronic pain dMRI studies. This diversity underscores a critical challenge for the field: the need for a more unified approach that would facilitate comparative and cumulative research. For a recent review of dMRI preprocessing, we refer to article ^131^and for a more detailed characterization of the impact of MRI acquisition parameters on diffusion models, we refer to international benchmark competitions reports, notably that discuss the impact of different inversion time (TI) and echo time (TE) ^132–134^.

### Processing and analysis tools

The tools provided by the FMRIB group at Oxford were the most used, with over 70% of the papers using one of the FSL tools to process their dMRI data. Almost 50% of the papers use the Tract Based Spatial Statistic (TBSS) pipeline ^135^ to identify voxel-wise differences. Although TBSS is a valid approach that provides significant advantages over classic whole-brain voxel-based analysis when it comes to group comparison, this method does not use the full potential of dMRI images as it summarizes the complexity of the whole-brain white matter into a WM skeleton that is only a few voxels wide. Also, this approach does not completely exclude registration errors ^136^. Therefore, almost no information coming from tracks spanning up to the cortex or tracks that are smaller or located in complex regions can be found in the TBSS maps. The other most commonly used software to process dMRI data were ExploreDTI ^137^ and MRtrix ^138^, both offering advanced tools specifically designed for dMRI data to generate tractography and diffusion metrics maps. To circumvent issues brought by TBSS, some studies presented a clever approach to identify white matter tracts impacted by the chronic pain condition under study. They used the clusters of significant differences identified in the TBSS results as seeds to perform probabilistic tractography. Although this approach allows for the identification of actual white matter tracts that were not present in the WM skeleton, it still cannot reconcile the fact that a significant volume of white matter was not included in the original TBSS analysis. Similarly, several groups identified GM ROIs from fMRI experiments and then expanded this region to include adjacent white matter as a WM ROI to extract diffusion properties. These approaches need to be interpreted carefully as a WM bundle passing close to a GM region does not necessarily connect with that region. Indeed, many WM tracts travel long distances in the brain without connecting with each region they are bordering along the way. A more appropriate approach could have been to use the GM ROI to generate seeds from/to which WM fibers might connect and extract anatomically plausible tracks from a whole brain tractogram.

To illustrate the constraints of the TBSS methods, we examined the disparities between TBSS and a track-based approach for two specific tracks, namely the fornix and the accumbofrontal (AcF) track, both of which are of particular significance in chronic pain research owing to the regions they connect. Notably, the fornix serves as the primary pathway for efferent signals from the hippocampus, and it plays a critical role in memory circuitry ^119^. Moreover, hippocampus volume has been shown to be a risk factor in the transition from acute to chronic pain ^34^. The AcF track connects the orbitofrontal cortex to the nucleus accumbens ^139^, both regions that are also implicated in the transition from acute to chronic pain ^30^. To demonstrate the benefits of employing targeted approaches for investigating white matter tracks involved in chronic pain, we analyzed the overlap of two streamline bundles extracted using separate techniques with a commonly used whole brain FA skeleton (method described in *supplementary material* [supp 3.]). As a quantitative measure of overlap, voxels intersected by these tracks were extracted and the percentage of voxels overlapping the binarized FA skeleton were output for each track. The fornix and AcF tracks are displayed in Figure 7 along with the percentage of overlapping voxels showing 15% voxels overlap between TBSS and tractography approached for the fornix (figure 7 left), and only 12% voxels overlap for the AcF track (figure 7 right). This analysis was reproduced for a few other subjects from the OpenPain database and similar overlaps were obtained (not shown).

**Figure 7.**
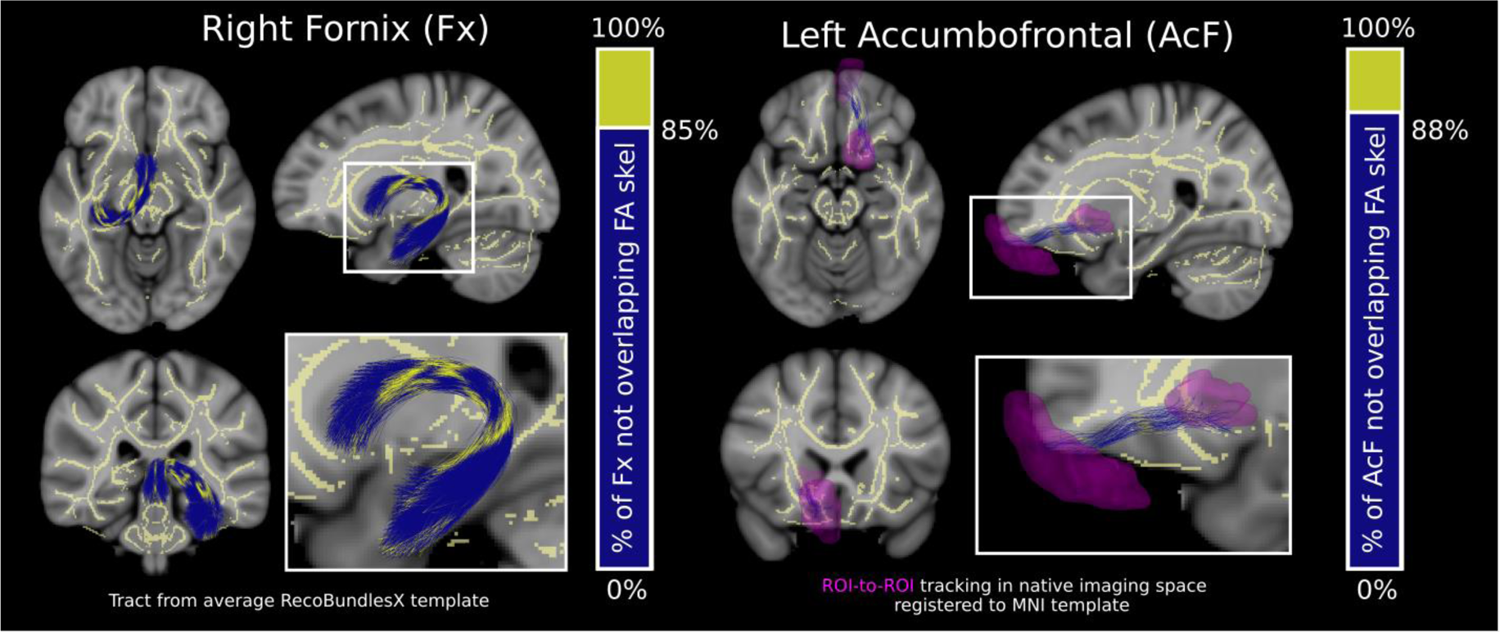
A visualization of the overlap between the FSL FA skeleton used commonly in whole brain diffusion MRI analysis and two different reconstructed white matter bundles taken from two different tractography analysis approaches. The population average right fornix (Fx) bundle used in the RecobundleX analysis pipeline is displayed in blue with regions overlapping the FA skeleton displayed in yellow. Voxels overlapping the bundle were extracted and the percentage of these voxels overlapping the FA skeleton are displayed as a bar graph. The same metrics are displayed for the Left Accumbofronal (AcF) track which was generated in native imaging space by extracting tracks that traversed ROIs (pink) from the nucleus accumbens and the orbital frontal gyrus. Only a small proportion of these tracks overlap the whole brain FA skeleton from FSL, suggesting that more targeted analysis approaches may be more sensitive to detecting microstructural alterations in some white matter structures.

### Where should the field go?

Similar to the challenges faced by fMRI for processing and analysis ^127,130^, dMRI is facing reproducibility and replication issues. As evidenced by the results of this review, there is a substantial degree of variability in published findings, methodologies, and metrics across different studies. While there exists several good guidelines discussing future directions for neuroimaging-based pain biomarker research ^3,25,140,141^, we suggest specific strategies to address dMRI’s challenges: i) updating dMRI signal modelling, ii) increase access and availability of dMRI data for CP, and iii) adopting standardized pipelines specific to dMRI.

**i) Updating dMRI local reconstruction methods:** To identify and increase group differences, novel biomarkers that increase sensitivity and specificity must be found. Recent advancements in magnetic resonance (MR) hardware and acquisition schemes— such as high angular resolution diffusion imaging (HARDI), diffusion spectrum imaging (DSI), and multi-shell protocols—as well as analysis methods enable the exploration of such brain markers ^142^. However, the field of chronic pain (CP) has yet to yield the full potential of these methods. For instance, diffusion tensor imaging (DTI) has been so prevalent that it is often used interchangeably with dMRI in literature. More recent studies in dMRI have moved away from DTI, as it fails to accurately represent multiple fiber populations within a single white matter voxel. The tensor model typically falls short in capturing accurate microstructural information in voxels where fiber crossing occurs; given that this happens in approximately 60 to 90% of all white matter voxels in the brain ^143^, the DTI method is increasingly seen as providing anatomically unsound information – especially when applying tractography. Recent dMRI research has begun to move away from DTI, adopting newer methods aiming for a more precise representation of underlying white matter tissue organization. Within the studies reviewed here, four studies employed whole-brain connectome metrics and two utilized multi-compartment local models. While these newer methods are promising, a caveat is warranted: if HARDI and multi-shell protocols gain widespread acceptance without standardization, there is a risk of exacerbating variability and noise in the CP literature. Notably, the application of these methods on DTI data will most likely provide unreliable results. Therefore, we advocate for the adoption of these advanced techniques only within a structured framework of standardized protocols, reproducible analytical pipelines and cross-validation.
**ii) Increasing access and availability of dMRI data for pain:** One of the notable challenges in the study of chronic pain using diffusion magnetic resonance imaging (dMRI) is the limited access and availability of extensive open-access datasets. These limitations hamper the statistical power and generalizability of research findings. As such, studies included in this review had a maximum of 103 participants and a minimum of 7 participants (with an average of 37.33 ± 27.39 participants). Recent advancements in the democratization of machine learning methods, in data sharing ^144,145^, in data harmonization ^146,147^, and the adoption of uniform metrics present significant opportunities for addressing this issue. By leveraging these advancements, researchers have the potential to rapidly expand and enrich open-access datasets specifically focused on chronic pain.
**iii) Adopting standardized pipelines specific to dMRI:** Both the field of chronic pain research and diffusion magnetic resonance imaging (dMRI) are subject to large intra-group variability. For chronic pain, such variability can be attenuated by refining the specificity of clinical evaluations, thereby reducing confounds related to the chronic pain condition itself. One approach could be selecting study participants with consistent clinical criteria or ensuring a large enough sample size for data-driven selection. For dMRI, variations can be minimized at every stage, from data collection to analysis. It is important to note that dMRI has specific challenges, unlike other types of structural MR imaging. These challenges include image susceptibility distortions, region-of-interest positioning, image registration, and data smoothing. Conventional structural MR analysis techniques, such as whole-brain voxel-based analysis, might be less suitable due to common dMRI pitfalls ^148^. Therefore, we recommend that the acquisition parameters, pre-processing, processing and analysis be conducted with standardized pipelines specific to dMRI.

Overall, the goal of this review was not to favor one method over another but to provide an overview of the current state of the field. However, in writing this review, we emphasize the difficulty of finding commonalities amidst the diverse methods used for image acquisition, analysis, and communication. Consequently, we pinpoint specific areas that require attention and potential improvements. Ultimately, we hope that addressing these challenges will allow pain researchers to capture more reproducible, specific and subtle white matter abnormalities.

## Limitations

Due to evolving chronic pain definitions and dMRI nomenclature, it is likely that articles performing dMRI on primary chronic pain patients were missed in this review ^48^. Some papers might have identified their participants otherwise had they used these new definitions. For example, an article from Geha et al ^117^, referenced by other studies in this review, was erroneously excluded because its dMRI nomenclature did not meet our inclusion criteria.

## Author Approval

All authors have seen and approved the manuscript.

## Conflits of interest

MD is co-founder of IMEKA inc.

All other authors declare no conflict of interest.

## Data Availability

All data produced in the present work are contained in the manuscript

https://osf.io/4wyqt/

## References

1. Baliki MN, Apkarian AV. Nociception, pain, negative moods and behavior selection. Neuron. 2015;87(3):474–491. doi:10.1016/j.neuron.2015.06.005

2. Kregel J, Meeus M, Malfliet A, et al. Structural and functional brain abnormalities in chronic low back pain: A systematic review. Semin Arthritis Rheum. 2015;45(2):229–237. doi:10.1016/j.semarthrit.2015.05.002

3. van der Miesen MM, Lindquist MA, Wager TD. Neuroimaging-based biomarkers for pain: state of the field and current directions. Pain Rep. 2019;4(4):e751. doi:10.1097/PR9.0000000000000751

4. Drake-Pérez M, Boto J, Fitsiori A, Lovblad K, Vargas MI. Clinical applications of diffusion weighted imaging in neuroradiology. Insights Imaging. 2018;9(4):535–547. doi:10.1007/s13244-018-0624-3

5. Tae WS, Ham BJ, Pyun SB, Kang SH, Kim BJ. Current Clinical Applications of Diffusion-Tensor Imaging in Neurological Disorders. *Journal of Clinical Neurology (Seoul*, Korea). 2018;14(2):129. doi:10.3988/jcn.2018.14.2.129

6. Nørhøj Jespersen S. White matter biomarkers from diffusion MRI. Journal of Magnetic Resonance. 2018;291:127–140. doi:10.1016/j.jmr.2018.03.001

7. Goveas J, O’Dwyer L, Mascalchi M, et al. Diffusion-MRI in neurodegenerative disorders. Magnetic Resonance Imaging. 2015;33(7):853–876. doi:10.1016/j.mri.2015.04.006

8. Kamagata K, Andica C, Kato A, et al. Diffusion Magnetic Resonance Imaging-Based Biomarkers for Neurodegenerative Diseases. Int J Mol Sci. 2021;22(10):5216. doi:10.3390/ijms22105216

9. Fields RD. Change in the Brain’s White Matter. Science. 2010;330(6005):768–769. doi:10.1126/science.1199139

10. Sharma J, Angelucci A, Sur M. Induction of visual orientation modules in auditory cortex. Nature. 2000;404(6780):841–847. doi:10.1038/35009043

11. Lebel C, Deoni S. The development of brain white matter microstructure. NeuroImage. 2018;182:207–218. doi:10.1016/j.neuroimage.2017.12.097

12. Kaiser M. Mechanisms of Connectome Development. Trends in Cognitive Sciences. 2017;21(9):703–717. doi:10.1016/j.tics.2017.05.010

13. The emergent properties of the connected brain | Science. Accessed November 7, 2023. https://www.science.org/doi/full/10.1126/science.abq2591

14. Forkel SJ, Friedrich P, Thiebaut de Schotten M, Howells H. White matter variability, cognition, and disorders: a systematic review. Brain Struct Funct. 2022;227(2):529–544. doi:10.1007/s00429-021-02382-w

15. Thiebaut de Schotten M, Foulon C, Nachev P. Brain disconnections link structural connectivity with function and behaviour. Nat Commun. 2020;11(1):5094. doi:10.1038/s41467-020-18920-9

16. Huber E, Donnelly PM, Rokem A, Yeatman JD. Rapid and widespread white matter plasticity during an intensive reading intervention. Nat Commun. 2018;9(1):2260. doi:10.1038/s41467-018-04627-5

17. Hutchinson EB, Schwerin SC, Avram AV, Juliano SL, Pierpaoli C. Diffusion MRI and the detection of alterations following traumatic brain injury. J Neurosci Res. 2018;96(4):612–625. doi:10.1002/jnr.24065

18. Andica C, Kamagata K, Hatano T, et al. MR Biomarkers of Degenerative Brain Disorders Derived From Diffusion Imaging. J Magn Reson Imaging. 2020;52(6):1620–1636. doi:10.1002/jmri.27019

19. Meijer FJA, Bloem BR, Mahlknecht P, Seppi K, Goraj B. Update on diffusion MRI in Parkinson’s disease and atypical parkinsonism. J Neurol Sci. 2013;332(1-2):21–29. doi:10.1016/j.jns.2013.06.032

20. Lakhani DA, Schilling KG, Xu J, Bagnato F. Advanced Multicompartment Diffusion MRI Models and Their Application in Multiple Sclerosis. AJNR Am J Neuroradiol. 2020;41(5):751–757. doi:10.3174/ajnr.A6484

21. Valdés Cabrera D, Stobbe R, Smyth P, Giuliani F, Emery D, Beaulieu C. Diffusion tensor imaging tractography reveals altered fornix in all diagnostic subtypes of multiple sclerosis. Brain Behav. 2019;10(1):e01514. doi:10.1002/brb3.1514

22. Wang L, Leonards CO, Sterzer P, Ebinger M. White matter lesions and depression: a systematic review and meta-analysis. J Psychiatr Res. 2014;56:56–64. doi:10.1016/j.jpsychires.2014.05.005

23. Tracey I, Woolf CJ, Andrews N. Composite pain biomarker signatures for objective assessment and effective treatment. Neuron. 2019;101(5):783–800. doi:10.1016/j.neuron.2019.02.019

24. Reckziegel D, Vachon-Presseau E, Petre B, Schnitzer TJ, Baliki MN, Apkarian AV. Deconstructing biomarkers for chronic pain: context- and hypothesis-dependent biomarker types in relation to chronic pain. Pain. 2019;160 Suppl 1(Suppl 1):S37–S48. doi:10.1097/j.pain.0000000000001529

25. Mackey S, Greely HT, Martucci KT. Neuroimaging-based pain biomarkers: definitions, clinical and research applications, and evaluation frameworks to achieve personalized pain medicine. Pain Rep. 2019;4(4):e762. doi:10.1097/PR9.0000000000000762

26. Woo CW, Chang LJ, Lindquist MA, Wager TD. Building better biomarkers: brain models in translational neuroimaging. Nat Neurosci. 2017;20(3):365–377. doi:10.1038/nn.4478

27. Mouraux A, Iannetti GD. The search for pain biomarkers in the human brain. Brain. 2018;141(12):3290–3307. doi:10.1093/brain/awy281

28. Tétreault P, Mansour A, Vachon-Presseau E, Schnitzer TJ, Apkarian AV, Baliki MN. Brain Connectivity Predicts Placebo Response across Chronic Pain Clinical Trials. PLOS Biology. 2016;14(10):e1002570. doi:10.1371/journal.pbio.1002570

29. Kutch JJ, Labus JS, Harris RE, et al. Resting-state functional connectivity predicts longitudinal pain symptom change in urologic chronic pelvic pain syndrome: a MAPP network study. PAIN. 2017;158(6):1069. doi:10.1097/j.pain.0000000000000886

30. Baliki MN, Petre B, Torbey S, et al. Corticostriatal functional connectivity predicts transition to chronic back pain. Nat Neurosci. 2012;15(8):1117–1119. doi:10.1038/nn.3153

31. Kutch JJ, Ichesco E, Hampson JP, et al. Brain signature and functional impact of centralized pain: a Multidisciplinary Approach to the Study of Chronic Pelvic Pain (MAPP) Network Study. Pain. 2017;158(10):1979–1991. doi:10.1097/j.pain.0000000000001001

32. Baliki MN, Schnitzer TJ, Bauer WR, Apkarian AV. Brain morphological signatures for chronic pain. PLoS One. 2011;6(10):e26010. doi:10.1371/journal.pone.0026010

33. Henn AT, Larsen B, Frahm L, et al. Structural imaging studies of patients with chronic pain: an anatomical likelihood estimate meta-analysis. Pain. 2023;164(1):e10–e24. doi:10.1097/j.pain.0000000000002681

34. Vachon-Presseau E, Tétreault P, Petre B, et al. Corticolimbic anatomical characteristics predetermine risk for chronic pain. Brain. 2016;139(7):1958–1970. doi:10.1093/brain/aww100

35. López-Solà M, Pujol J, Wager TD, et al. Altered Functional Magnetic Resonance Imaging Responses to Nonpainful Sensory Stimulation in Fibromyalgia Patients. Arthritis & Rheumatology. 2014;66(11):3200–3209. doi:10.1002/art.38781

36. Harte SE, Ichesco E, Hampson JP, et al. Pharmacologic attenuation of cross-modal sensory augmentation within the chronic pain insula. Pain. 2016;157(9):1933–1945. doi:10.1097/j.pain.0000000000000593

37. López-Solà M, Woo CW, Pujol J, et al. Towards a neurophysiological signature for fibromyalgia. Pain. 2017;158(1):34–47. doi:10.1097/j.pain.0000000000000707

38. Baliki MN, Geha PY, Fields HL, Apkarian AV. Predicting Value of Pain and Analgesia: Nucleus Accumbens Response to Noxious Stimuli Changes in the Presence of Chronic Pain.

39. Baliki MN, Mansour AR, Baria AT, Apkarian AV. Functional Reorganization of the Default Mode Network across Chronic Pain Conditions. PLoS One. 2014;9(9):e106133. doi:10.1371/journal.pone.0106133

40. Cheng JC, Rogachov A, Hemington KS, et al. Multivariate machine learning distinguishes cross-network dynamic functional connectivity patterns in state and trait neuropathic pain. PAIN. 2018;159(9):1764. doi:10.1097/j.pain.0000000000001264

41. Seminowicz DA, Wideman TH, Naso L, et al. Effective Treatment of Chronic Low Back Pain in Humans Reverses Abnormal Brain Anatomy and Function. J Neurosci. 2011;31(20):7540–7550. doi:10.1523/JNEUROSCI.5280-10.2011

42. Tétreault P, Baliki MN, Baria AT, Bauer WR, Schnitzer TJ, Apkarian AV. Inferring distinct mechanisms in the absence of subjective differences: Placebo and centrally acting analgesic underlie unique brain adaptations. Hum Brain Mapp. 2018;39(5):2210–2223. doi:10.1002/hbm.23999

43. Stejskal EO, Tanner JE. Spin Diffusion Measurements: Spin Echoes in the Presence of a Time-Dependent Field Gradient. The Journal of Chemical Physics. 2004;42(1):288–292. doi:10.1063/1.1695690

44. Tournier JD. Diffusion MRI in the brain – Theory and concepts. Progress in Nuclear Magnetic Resonance Spectroscopy. 2019;112–113:1-16. doi:10.1016/j.pnmrs.2019.03.001

45. Moseley ME, Cohen Y, Mintorovitch J, et al. Early detection of regional cerebral ischemia in cats: Comparison of diffusion- and T2-weighted MRI and spectroscopy. Magnetic Resonance in Medicine. 1990;14(2):330–346. doi:10.1002/mrm.1910140218

46. Basser PJ, Mattiello J, LeBihan D. MR diffusion tensor spectroscopy and imaging. Biophysical Journal. 1994;66(1):259–267. doi:10.1016/S0006-3495(94)80775-1

47. Tournier JD, Mori S, Leemans A. Diffusion tensor imaging and beyond. Magn Reson Med. 2011;65(6):1532–1556. doi:10.1002/mrm.22924

48. Nicholas M, Vlaeyen JWS, Rief W, et al. The IASP classification of chronic pain for ICD-11. Pain. 2019;160(1):28–37. doi:10.1097/j.pain.0000000000001390

49. Treede RD, Rief W, Barke A, et al. Chronic pain as a symptom or a disease: the IASP Classification of Chronic Pain for the International Classification of Diseases (ICD-11). Pain. 2019;160(1):19–27. doi:10.1097/j.pain.0000000000001384

50. Tournier JD, Yeh CH, Calamante F, Cho KH, Connelly A, Lin CP. Resolving crossing fibres using constrained spherical deconvolution: Validation using diffusion-weighted imaging phantom data. NeuroImage. 2008;42(2):617–625. doi:10.1016/j.neuroimage.2008.05.002

51. Zhang H, Schneider T, Wheeler-Kingshott CA, Alexander DC. NODDI: Practical in vivo neurite orientation dispersion and density imaging of the human brain. NeuroImage. 2012;61(4):1000–1016. doi:10.1016/j.neuroimage.2012.03.072

52. Smith SM, Jenkinson M, Woolrich MW, et al. Advances in functional and structural MR image analysis and implementation as FSL. NeuroImage. 2004;23:S208–S219. doi:10.1016/j.neuroimage.2004.07.051

53. Smith SM, Jenkinson M, Johansen-Berg H, et al. Tract-based spatial statistics: Voxelwise analysis of multi-subject diffusion data. NeuroImage. 2006;31(4):1487–1505. doi:10.1016/j.neuroimage.2006.02.024

54. O’Donnell LJ, Westin CF, Golby AJ. Tract-based morphometry for white matter group analysis. NeuroImage. 2009;45(3):832–844. doi:10.1016/j.neuroimage.2008.12.023

55. Yeatman JD, Dougherty RF, Myall NJ, Wandell BA, Feldman HM. Tract Profiles of White Matter Properties: Automating Fiber-Tract Quantification. PLOS ONE. 2012;7(11):e49790. doi:10.1371/journal.pone.0049790

56. Zalesky A, Fornito A, Bullmore ET. Network-based statistic: Identifying differences in brain networks. NeuroImage. 2010;53(4):1197–1207. doi:10.1016/j.neuroimage.2010.06.041

57. Serin E, Zalesky A, Matory A, Walter H, Kruschwitz JD. NBS-Predict: A prediction-based extension of the network-based statistic. Neuroimage. 2021;244:118625. doi:10.1016/j.neuroimage.2021.118625

58. Kattem Husøy A, Eikenes L, Håberg AK, Hagen K, Stovner LJ. Diffusion tensor imaging in middle-aged headache sufferers in the general population: a cross-sectional population-based imaging study in the Nord-Trøndelag health study (HUNT-MRI). J Headache Pain. 2019;20(1):78. doi:10.1186/s10194-019-1028-6

59. Zhang J, Wu YL, Su J, et al. Assessment of gray and white matter structural alterations in migraineurs without aura. J Headache Pain. 2017;18(1):74. doi:10.1186/s10194-017-0783-5

60. Chong CD, Berisha V, Ross K, Kahn M, Dumkrieger G, Schwedt TJ. Distinguishing persistent post-traumatic headache from migraine: Classification based on clinical symptoms and brain structural MRI data. Cephalalgia. Published online April 29, 2021:333102421991819. doi:10.1177/0333102421991819

61. Marciszewski KK, Meylakh N, Di Pietro F, Macefield VG, Macey PM, Henderson LA. Fluctuating Regional Brainstem Diffusion Imaging Measures of Microstructure across the Migraine Cycle. eNeuro. 2019;6(4). doi:10.1523/ENEURO.0005-19.2019

62. DaSilva AFM, Granziera C, Tuch DS, Snyder J, Vincent M, Hadjikhani N. Interictal alterations of the trigeminal somatosensory pathway and periaqueductal gray matter in migraine. Neuroreport. 2007;18(4):301–305. doi:10.1097/WNR.0b013e32801776bb

63. Planchuelo-Gómez Á, García-Azorín D, Guerrero ÁL, Aja-Fernández S, Rodríguez M, de Luis-García R. Multimodal fusion analysis of structural connectivity and gray matter morphology in migraine. Human Brain Mapping. 2021;42(4):908–921. doi:10.1002/hbm.25267

64. Neeb L, Bastian K, Villringer K, et al. No microstructural white matter alterations in chronic and episodic migraineurs: a case-control diffusion tensor magnetic resonance imaging study. Headache. 2015;55(2):241–251. doi:10.1111/head.12496

65. Coppola G, Di Renzo A, Tinelli E, et al. Patients with chronic migraine without history of medication overuse are characterized by a peculiar white matter fiber bundle profile. J Headache Pain. 2020;21(1):92. doi:10.1186/s10194-020-01159-6

66. Gomez-Beldarrain M, Oroz I, Zapirain BG, et al. Right fronto-insular white matter tracts link cognitive reserve and pain in migraine patients. J Headache Pain. 2015;17:4. doi:10.1186/s10194-016-0593-1

67. Szabó N, Kincses ZT, Párdutz Á, et al. White matter microstructural alterations in migraine: a diffusion-weighted MRI study. Pain. 2012;153(3):651–656. doi:10.1016/j.pain.2011.11.029

68. Silvestro M, Tessitore A, Caiazzo G, et al. Disconnectome of the migraine brain: a “connectopathy” model. J Headache Pain. 2021;22(1):102. doi:10.1186/s10194-021-01315-6

69. Szabó N, Kincses ZT, Párdutz A, et al. White matter disintegration in cluster headache. The journal of headache and pain. 2013;14:64. doi:10.1186/1129-2377-14-64

70. Teepker M, Menzler K, Belke M, et al. Diffusion tensor imaging in episodic cluster headache. Headache. 2012;52(2):274–282. doi:10.1111/j.1526-4610.2011.02000.x

71. Miller JV, Andre Q, Timmers I, et al. Subclinical post-traumatic stress symptomology and brain structure in youth with chronic headaches. Neuroimage Clin. 2021;30:102627. doi:10.1016/j.nicl.2021.102627

72. Salomons TV, Moayedi M, Weissman-Fogel I, et al. Perceived helplessness is associated with individual differences in the central motor output system. Eur J Neurosci. 2012;35(9):1481–1487. doi:10.1111/j.1460-9568.2012.08048.x

73. Moayedi M, Weissman-Fogel I, Salomons TV, et al. White matter brain and trigeminal nerve abnormalities in temporomandibular disorder. Pain. 2012;153(7):1467–1477. doi:10.1016/j.pain.2012.04.003

74. Gustin SM, Peck CC, Cheney LB, Macey PM, Murray GM, Henderson LA. Pain and plasticity: is chronic pain always associated with somatosensory cortex activity and reorganization? J Neurosci. 2012;32(43):14874–14884. doi:10.1523/JNEUROSCI.1733-12.2012

75. Wada A, Shizukuishi T, Kikuta J, et al. Altered structural connectivity of pain-related brain network in burning mouth syndrome-investigation by graph analysis of probabilistic tractography. Neuroradiology. 2017;59(5):525–532. doi:10.1007/s00234-017-1830-2

76. Khan SA, Keaser ML, Meiller TF, Seminowicz DA. Altered structure and function in the hippocampus and medial prefrontal cortex in patients with burning mouth syndrome. Pain. 2014;155(8):1472–1480. doi:10.1016/j.pain.2014.04.022

77. Tan Y, Wu X, Chen J, Kong L, Qian Z. Structural and Functional Connectivity Between the Amygdala and Orbital Frontal Cortex in Burning Mouth Syndrome: An fMRI Study. Front Psychol. 2019;10:1700. doi:10.3389/fpsyg.2019.01700

78. Kurokawa R, Kamiya K, Inui S, et al. Structural connectivity changes in the cerebral pain matrix in burning mouth syndrome: a multi-shell, multi-tissue-constrained spherical deconvolution model analysis. Neuroradiology. 2021;63(12):2005–2012. doi:10.1007/s00234-021-02732-9

79. Dun W, Yang J, Yang L, et al. Abnormal white matter integrity during pain-free periovulation is associated with pain intensity in primary dysmenorrhea. Brain Imaging Behav. 2017;11(4):1061–1070. doi:10.1007/s11682-016-9582-x

80. He J, Dun W, Han F, et al. Abnormal white matter microstructure along the thalamus fiber pathways in women with primary dysmenorrhea. Brain Imaging and Behavior. Published online 2020. doi:10.1007/s11682-020-00400-9

81. Liu J, Liu H, Mu J, et al. Altered white matter microarchitecture in the cingulum bundle in women with primary dysmenorrhea: A tract-based analysis study. Hum Brain Mapp. 2017;38(9):4430–4443. doi:10.1002/hbm.23670

82. Liu P, Wang G, Liu Y, et al. White matter microstructure alterations in primary dysmenorrhea assessed by diffusion tensor imaging. Sci Rep. 2016;6:25836. doi:10.1038/srep25836

83. Irimia A, Labus JS, Torgerson CM, Van Horn JD, Mayer EA. Altered viscerotopic cortical innervation in patients with irritable bowel syndrome. Neurogastroenterol Motil. 2015;27(8):1075–1081. doi:10.1111/nmo.12586

84. Ellingson BM, Mayer E, Harris RJ, et al. Diffusion tensor imaging detects microstructural reorganization in the brain associated with chronic irritable bowel syndrome. Pain. 2013;154(9):1528–1541. doi:10.1016/j.pain.2013.04.010

85. Qi R, Liu C, Weng Y, et al. Disturbed Interhemispheric Functional Connectivity Rather than Structural Connectivity in Irritable Bowel Syndrome. Front Mol Neurosci. 2016;9:141. doi:10.3389/fnmol.2016.00141

86. Hubbard CS, Becerra L, Heinz N, et al. Microstructural White Matter Abnormalities in the Dorsal Cingulum of Adolescents with IBS. eNeuro. 2018;5(4). doi:10.1523/ENEURO.0354-17.2018

87. Chen JYW, Blankstein U, Diamant NE, Davis KD. White matter abnormalities in irritable bowel syndrome and relation to individual factors. Brain Res. 2011;1392:121–131. doi:10.1016/j.brainres.2011.03.069

88. Nan J, Zhang L, Chen Q, et al. White Matter Microstructural Similarity and Diversity of Functional Constipation and Constipation-predominant Irritable Bowel Syndrome. J Neurogastroenterol Motil. 2018;24(1):107–118. doi:10.5056/jnm17038

89. Liu G, Li S, Chen N, et al. Inter-hemispheric Functional Connections Are More Vulnerable to Attack Than Structural Connection in Patients With Irritable Bowel Syndrome. J Neurogastroenterol Motil. 2021;27(3):426–435. doi:10.5056/jnm20134

90. Woodworth D, Mayer E, Leu K, et al. Unique Microstructural Changes in the Brain Associated with Urological Chronic Pelvic Pain Syndrome (UCPPS) Revealed by Diffusion Tensor MRI, Super-Resolution Track Density Imaging, and Statistical Parameter Mapping: A MAPP Network Neuroimaging Study. PLoS One. 2015;10(10):e0140250. doi:10.1371/journal.pone.0140250

91. Huang L, Kutch JJ, Ellingson BM, et al. Brain white matter changes associated with urological chronic pelvic pain syndrome: multisite neuroimaging from a MAPP case-control study. Pain. 2016;157(12):2782–2791. doi:10.1097/j.pain.0000000000000703

92. Woodworth DC, Dagher A, Curatolo A, et al. Changes in brain white matter structure are associated with urine proteins in urologic chronic pelvic pain syndrome (UCPPS): A MAPP Network study. PLoS One. 2018;13(12):e0206807. doi:10.1371/journal.pone.0206807

93. Alger JR, Ellingson BM, Ashe-McNalley C, et al. Multisite, multimodal neuroimaging of chronic urological pelvic pain: Methodology of the MAPP Research Network. Neuroimage Clin. 2016;12:65–77. doi:10.1016/j.nicl.2015.12.009

94. Farmer MA, Chanda ML, Parks EL, Baliki MN, Apkarian AV, Schaeffer AJ. Brain functional and anatomical changes in chronic prostatitis/chronic pelvic pain syndrome. J Urol. 2011;186(1):117–124. doi:10.1016/j.juro.2011.03.027

95. Huang X, Chen J, Liu S, et al. Impaired frontal-parietal control network in chronic prostatitis/chronic pelvic pain syndrome revealed by graph theoretical analysis: A DTI study. Eur J Neurosci. 2021;53(4):1060–1071. doi:10.1111/ejn.14962

96. Farmer MA, Huang L, Martucci K, et al. Brain White Matter Abnormalities in Female Interstitial Cystitis/Bladder Pain Syndrome: A MAPP Network Neuroimaging Study. J Urol. 2015;194(1):118–126. doi:10.1016/j.juro.2015.02.082

97. Gupta A, Woodworth DC, Ellingson BM, et al. Disease-Related Microstructural Differences in the Brain in Women With Provoked Vestibulodynia. J Pain. 2018;19(5):528.e1-528.e15. doi:10.1016/j.jpain.2017.12.269

98. Mansour AR, Baliki MN, Huang L, et al. Brain white matter structural properties predict transition to chronic pain. Pain. 2013;154(10):2160–2168. doi:10.1016/j.pain.2013.06.044

99. Ma J, Wang X, Qiu Q, Zhan H, Wu W. Changes in Empathy in Patients With Chronic Low Back Pain: A Structural-Functional Magnetic Resonance Imaging Study. Front Hum Neurosci. 2020;14:326. doi:10.3389/fnhum.2020.00326

100. Čeko M, Shir Y, Ouellet JA, Ware MA, Stone LS, Seminowicz DA. Partial recovery of abnormal insula and dorsolateral prefrontal connectivity to cognitive networks in chronic low back pain after treatment. Hum Brain Mapp. 2015;36(6):2075–2092. doi:10.1002/hbm.22757

101. Kim H, Mawla I, Lee J, et al. Reduced tactile acuity in chronic low back pain is linked with structural neuroplasticity in primary somatosensory cortex and is modulated by acupuncture therapy. Neuroimage. 2020;217:116899. doi:10.1016/j.neuroimage.2020.116899

102. Bishop JH, Shpaner M, Kubicki A, Clements S, Watts R, Naylor MR. Structural network differences in chronic muskuloskeletal pain: Beyond fractional anisotropy. Neuroimage. 2018;182:441–455. doi:10.1016/j.neuroimage.2017.12.021

103. Lieberman G, Shpaner M, Watts R, et al. White matter involvement in chronic musculoskeletal pain. J Pain. 2014;15(11):1110–1119. doi:10.1016/j.jpain.2014.08.002

104. Cruz-Almeida Y, Coombes S, Febo M. Pain Differences in Neurite Orientation and Dispersion Density Imaging Measures Among Community-Dwelling Older Adults. Exp Gerontol. 2021;154:111520. doi:10.1016/j.exger.2021.111520

105. Pijnenburg M, Hosseini SMH, Brumagne S, Janssens L, Goossens N, Caeyenberghs K. Structural Brain Connectivity and the Sit-to-Stand-to-Sit Performance in Individuals with Nonspecific Low Back Pain: A Diffusion Magnetic Resonance Imaging-Based Network Analysis. Brain Connect. 2016;6(10):795–803. doi:10.1089/brain.2015.0401

106. Coppieters I, De Pauw R, Caeyenberghs K, et al. Differences in white matter structure and cortical thickness between patients with traumatic and idiopathic chronic neck pain: Associations with cognition and pain modulation? Hum Brain Mapp. 2018;39(4):1721–1742. doi:10.1002/hbm.23947

107. Butler S, Landmark T, Glette M, Borchgrevink P, Woodhouse A. Chronic widespread pain-the need for a standard definition. Pain. 2016;157(3):541–543. doi:10.1097/j.pain.0000000000000417

108. Sundgren PC, Petrou M, Harris RE, et al. Diffusion-weighted and diffusion tensor imaging in fibromyalgia patients: a prospective study of whole brain diffusivity, apparent diffusion coefficient, and fraction anisotropy in different regions of the brain and correlation with symptom severity. Acad Radiol. 2007;14(7):839–846. doi:10.1016/j.acra.2007.03.015

109. Ceko M, Bushnell MC, Fitzcharles MA, Schweinhardt P. Fibromyalgia interacts with age to change the brain. Neuroimage Clin. 2013;3:249–260. doi:10.1016/j.nicl.2013.08.015

110. Kim H, Kim J, Loggia ML, et al. Fibromyalgia is characterized by altered frontal and cerebellar structural covariance brain networks. Neuroimage Clin. 2015;7:667–677. doi:10.1016/j.nicl.2015.02.022

111. Hadanny A, Bechor Y, Catalogna M, et al. Hyperbaric Oxygen Therapy Can Induce Neuroplasticity and Significant Clinical Improvement in Patients Suffering From Fibromyalgia With a History of Childhood Sexual Abuse-Randomized Controlled Trial. Front Psychol. 2018;9:2495. doi:10.3389/fpsyg.2018.02495

112. Fayed N, Garcia-Campayo J, Magallón R, et al. Localized 1H-NMR spectroscopy in patients with fibromyalgia: a controlled study of changes in cerebral glutamate/glutamine, inositol, choline, and N-acetylaspartate. Arthritis Res Ther. 2010;12(4):R134. doi:10.1186/ar3072

113. Lutz J, Jäger L, de Quervain D, et al. White and gray matter abnormalities in the brain of patients with fibromyalgia: a diffusion-tensor and volumetric imaging study. Arthritis Rheum. 2008;58(12):3960–3969. doi:10.1002/art.24070

114. Birklein F, Schlereth T. Complex regional pain syndrome-significant progress in understanding. Pain. 2015;156 Suppl 1:S94–S103. doi:10.1097/01.j.pain.0000460344.54470.20

115. Bruehl S. Complex regional pain syndrome. BMJ. 2015;351:h2730. doi:10.1136/bmj.h2730

116. Hotta J, Zhou G, Harno H, Forss N, Hari R. Complex regional pain syndrome: The matter of white matter? Brain Behav. 2017;7(5):e00647. doi:10.1002/brb3.647

117. Geha PY, Baliki MN, Harden RN, Bauer WR, Parrish TB, Apkarian AV. The brain in chronic CRPS pain: Abnormal gray-white matter interactions in emotional and autonomic regions. Neuron. 2008;60(4):570–581. doi:10.1016/j.neuron.2008.08.022

118. Jennings RG, Van Horn JD. Publication bias in neuroimaging research: Implications for meta-analyses. Neuroinformatics. 2012;10(1):67–80. doi:10.1007/s12021-011-9125-y

119. Cahn AJ, Little G, Beaulieu C, Tétreault P. Diffusion properties of the fornix assessed by deterministic tractography shows age, sex, volume, cognitive, hemispheric, and twin relationships in young adults from the Human Connectome Project. Brain Struct Funct. 2021;226(2):381–395. doi:10.1007/s00429-020-02181-9

120. Cai LY, Yang Q, Kanakaraj P, et al. MASiVar: Multisite, multiscanner, and multisubject acquisitions for studying variability in diffusion weighted MRI. Magn Reson Med. 2021;86(6):3304–3320. doi:10.1002/mrm.28926

121. Burdette JH, Durden DD, Elster AD, Yen YF. High b-value diffusion-weighted MRI of normal brain. J Comput Assist Tomogr. 2001;25(4):515–519. doi:10.1097/00004728-200107000-00002

122. Lebel C, Gee M, Camicioli R, Wieler M, Martin W, Beaulieu C. Diffusion tensor imaging of white matter tract evolution over the lifespan. Neuroimage. 2012;60(1):340–352. doi:10.1016/j.neuroimage.2011.11.094

123. Ni H, Kavcic V, Zhu T, Ekholm S, Zhong J. Effects of Number of Diffusion Gradient Directions on Derived Diffusion Tensor Imaging Indices in Human Brain. AJNR Am J Neuroradiol. 2006;27(8):1776–1781.

124. Oouchi H, Yamada K, Sakai K, et al. Diffusion Anisotropy Measurement of Brain White Matter Is Affected by Voxel Size: Underestimation Occurs in Areas with Crossing Fibers. AJNR Am J Neuroradiol. 2007;28(6):1102–1106. doi:10.3174/ajnr.A0488

125. Wu YC, Alexander AL. Hybrid diffusion imaging. Neuroimage. 2007;36(3):617–629. doi:10.1016/j.neuroimage.2007.02.050

126. Basser PJ, Mattiello J, LeBihan D. Estimation of the effective self-diffusion tensor from the NMR spin echo. J Magn Reson B. 1994;103(3):247–254. doi:10.1006/jmrb.1994.1037

127. Shrager RI, Basser PJ. Anisotropically weighted MRI. Magn Reson Med. 1998; 40(1):160–165. doi:10.1002/mrm.1910400121

128. Jeurissen B, Descoteaux M, Mori S, Leemans A. Diffusion MRI fiber tractography of the brain. NMR in Biomedicine. 2019;32(4):e3785. doi:10.1002/nbm.3785

129. Dell’Acqua F, Tournier JD. Modelling white matter with spherical deconvolution: How and why? NMR in Biomedicine. 2019;32(4):e3945. doi:10.1002/nbm.3945

130. Neher P, Stieltjes B, Wolf I, Meinzer HP, Maier-Hein K. Analysis of tractography biases introduced by anisotropic voxels. In:; 2013.

131. Tax CMW, Bastiani M, Veraart J, Garyfallidis E, Okan Irfanoglu M. What’s new and what’s next in diffusion MRI preprocessing. Neuroimage. 2022;249:118830. doi:10.1016/j.neuroimage.2021.118830

132. De Luca A, Ianus A, Leemans A, et al. On the generalizability of diffusion MRI signal representations across acquisition parameters, sequences and tissue types: Chronicles of the MEMENTO challenge. NeuroImage. 2021;240:118367. doi:10.1016/j.neuroimage.2021.118367

133. Diffusion MRI microstructure models with in vivo human brain Connectome data: results from a multi-group comparison - Ferizi - 2017 - NMR in Biomedicine - Wiley Online Library. Accessed December 14, 2023. https://analyticalsciencejournals.onlinelibrary.wiley.com/doi/10.1002/nbm.3734

134. Pizzolato M, Palombo M, Bonet-Carne E, et al. Acquiring and Predicting Multidimensional Diffusion (MUDI) Data: An Open Challenge. In: Bonet-Carne E, Hutter J, Palombo M, Pizzolato M, Sepehrband F, Zhang F, eds. Computational Diffusion MRI. Mathematics and Visualization. Springer International Publishing; 2020:195–208. doi:10.1007/978-3-030-52893-5_17

135. Smith SM, Jenkinson M, Johansen-Berg H, et al. Tract-based spatial statistics: voxelwise analysis of multi-subject diffusion data. Neuroimage. 2006;31(4):1487–1505. doi:10.1016/j.neuroimage.2006.02.024

136. Bach M, Laun FB, Leemans A, et al. Methodological considerations on tract-based spatial statistics (TBSS). NeuroImage. 2014;100:358–369. doi:10.1016/j.neuroimage.2014.06.021

137. Leemans A, Jeurissen B, Sijbers J, Jones DK. ExploreDTI: A graphical toolbox for processing, analyzing, and visualizing diffusion MR data. 17th Annual Meeting of Intl Soc Mag Reson Med. 2009;17.

138. Tournier JD, Smith R, Raffelt D, et al. MRtrix3: A fast, flexible and open software framework for medical image processing and visualisation. Neuroimage. 2019;202:116137. doi:10.1016/j.neuroimage.2019.116137

139. Karlsgodt KH, John M, Ikuta T, et al. The accumbofrontal tract: Diffusion tensor imaging characterization and developmental change from childhood to adulthood. Human Brain Mapping. 2015;36(12):4954. doi:10.1002/hbm.22989

140. Davis KD, Aghaeepour N, Ahn AH, et al. Discovery and validation of biomarkers to aid the development of safe and effective pain therapeutics: challenges and opportunities. Nat Rev Neurol. 2020;16(7):381–400. doi:10.1038/s41582-020-0362-2

141. Sluka KA, Wager TD, Sutherland SP, et al. Predicting chronic postsurgical pain: current evidence and a novel program to develop predictive biomarker signatures. PAIN. 2023;164(9):1912. doi:10.1097/j.pain.0000000000002938

142. Assaf Y, Johansen-Berg H, Thiebaut de Schotten M. The role of diffusion MRI in neuroscience. NMR Biomed. 2019;32(4):e3762. doi:10.1002/nbm.3762

143. Jeurissen B, Leemans A, Tournier J, Jones DK, Sijbers J. Investigating the prevalence of complex fiber configurations in white matter tissue with diffusion magnetic resonance imaging. Hum Brain Mapp. 2013;34(11):2747–2766. doi:10.1002/hbm.22099

144. Gorgolewski KJ, Auer T, Calhoun VD, et al. The brain imaging data structure, a format for organizing and describing outputs of neuroimaging experiments. Sci Data. 2016;3(1):160044. doi:10.1038/sdata.2016.44

145. Gorgolewski KJ, Varoquaux G, Rivera G, et al. NeuroVault.org: A repository for sharing unthresholded statistical maps, parcellations, and atlases of the human brain. NeuroImage. 2016;124:1242–1244. doi:10.1016/j.neuroimage.2015.04.016

146. Orlhac F, Eertink JJ, Cottereau AS, et al. A Guide to ComBat Harmonization of Imaging Biomarkers in Multicenter Studies. J Nucl Med. 2022;63(2):172–179. doi:10.2967/jnumed.121.262464

147. Mirzaalian H, de Pierrefeu A, Savadjiev P, et al. Harmonizing Diffusion MRI Data Across Multiple Sites and Scanners. Med Image Comput Comput Assist Interv. 2015;9349:12–19. doi:10.1007/978-3-319-24553-9_2

148. Jones DK, Cercignani M. Twenty-five pitfalls in the analysis of diffusion MRI data. NMR Biomed. 2010;23(7):803–820. doi:10.1002/nbm.1543

