## Supplementary material for "What has brain diffusion MRI taught us about chronic pain: a narrative review"

**Supp 1. Search strategies**

In PubMed, the following strategy has been used:

((("mri"[Title/Abstract] OR "magnetic resonance imaging"[Title/Abstract] OR "NMR"[Title/Abstract] OR "nuclear magnetic resonance"[Title/Abstract]) AND ("diffusion"[Title/Abstract] OR "DTI"[Title/Abstract])) OR "tensor imaging"[Title/Abstract]) AND "pain"[Title/Abstract] AND "brain"[Title/Abstract]

In PubMed (MeSH), the following strategy has been used:

"Diffusion Magnetic Resonance Imaging"[MeSH Terms] AND "Pain"[MeSH Terms] AND "Brain"[MeSH Terms]

In Scopus, the follow strategy has been used:

( TITLE-ABS-KEY ( ( ( mri OR "magnetic resonance imaging" OR nmr OR "nuclear magnetic resonance" ) W/2 ( diffusion ) ) OR dti OR "tensor imaging" ) AND TITLE-ABS-KEY ( pain ) AND TITLE-ABS-KEY ( brain ) ) AND NOT INDEX ( medline )

**Supp 2. A note on acute pain**

The link between white matter characteristics and acute pain perception, as often reported by pain thermal stimulation fMRI studies, is under-studied. Preliminary screening revealed that only 13 studies specifically investigated this aspect. Considering the significant methodological differences compared to chronic pain, acute pain studies were excluded from this review.

**Supp 3. Method description for figure 7**

For the first technique, data from a single subject from Mansour et coll. paper [1], that can be found on the OpenPain neuroimaging dataset platform (https://openpain.org) was used to generate a track segmentation of the left AcF track in native imaging space. Firstly, the anatomical (T1w structural image, 1 mm isotropic) and diffusion data (8 b0 s/mm$^2$, 60 b1000 s/mm$^2$, whole brain 2 mm isotropic) were processed using tractoflow version 2.3.0 [2] outputting a particle filtered whole brain tractogram [3]. Next the Brainnetome atlas [4] was registered to the subject space using the T1w image and a non-linear registration using the ANTS toolbox [5] followed by a nearest neighbor interpolation of the Brainnetome atlas to the diffusion data. Streamlines for the left AcF were segmented by selecting all streamlines that joined the brainnetome ROIs (left nucleus accumbens and the left orbital frontal gyrus). The AcF track was then registered to MNI space using the previously calculated non-linear transformation. For comparison of the FA skeleton to multiple methods, the population averaged right fornix used in the RecobundlesX analysis pipeline [6–9] along with the aforementioned left AcF track were overlaid on the binarized whole brain FA skeleton.

**Supp references**


1. Mansour AR, Baliki MN, Huang L, et al. Brain white matter structural properties predict transition to chronic pain. *Pain*. 2013;154(10):2160-2168. doi:10.1016/j.pain.2013.06.044

2. Theaud G, Houde JC, Boré A, Rheault F, Morency F, Descoteaux M. TractoFlow: A robust, efficient and reproducible diffusion MRI pipeline leveraging Nextflow & Singularity. *NeuroImage*. 2020;218:116889. doi:10.1016/j.neuroimage.2020.116889

3. Girard G, Whittingstall K, Deriche R, Descoteaux M. Towards quantitative connectivity analysis: reducing tractography biases. *Neuroimage*. 2014;98:266-278. doi:10.1016/j.neuroimage.2014.04.074

4. Fan L, Li H, Zhuo J, et al. The Human Brainnetome Atlas: A New Brain Atlas Based on Connectional Architecture. *Cereb Cortex*. 2016;26(8):3508-3526. doi:10.1093/cercor/bhw157

5. Avants BB, Tustison NJ, Stauffer M, Song G, Wu B, Gee JC. The Insight ToolKit image registration framework. *Front Neuroinform*. 2014;8:44. doi:10.3389/fninf.2014.00044

6. St-Onge E, Schilling K, Rheault F. *BundleSeg: A Versatile, Reliable and Reproducible Approach to White Matter Bundle Segmentation.*; 2023.

7. Kurtzer GM, Sochat V, Bauer MW. Singularity: Scientific containers for mobility of compute. *PLOS ONE*. 2017;12(5):e0177459. doi:10.1371/journal.pone.0177459

8. Di Tommaso P, Chatzou M, Floden EW, Barja PP, Palumbo E, Notredame C. Nextflow enables reproducible computational workflows. *Nat Biotechnol*. 2017;35(4):316-319. doi:10.1038/nbt.3820

9. Rheault F. Analyse et reconstruction de faisceaux de la matière blanche. Published online 2020. Accessed December 21, 2023. https://savoirs.usherbrooke.ca/handle/11143/17255
